# *GRID1/*GluD1 homozygous variants linked to intellectual disability and spastic paraplegia impair mGlu1/5 receptor signaling and excitatory synapses

**DOI:** 10.1101/2022.05.16.22274994

**Authors:** Dévina C. Ung, Ludovic Tricoire, Nicolas Pietrancosta, Andjela Zlatanovic, Ben Pode-Shakked, Annick Raas-Rothschild, Orly Elpeleg, Bassam Abu-Libdeh, Nasrin Hamed, Marie-Amélie Papon, Sylviane Marouillat, Rose-Anne Thépault, Giovanni Stevanin, Bertrand Lambolez, Annick Toutain, Régine Hepp, Frédéric Laumonnier

**Author notes:** These authors contributed equally to this work (Bertrand Lambolez, Régine Hepp, Frédéric Laumonnier). **<u>Address for correspondence:</u>** - Dr Régine HEPP, PhD, Sorbonne Université, Neuroscience Paris Seine, 9 quai St Bernard, 75005 Paris France., - Dr Frédéric LAUMONNIER, PhD, UMR1253 INSERM; Team neurogenomics and neuronal physiopathology; Faculty of Medicine; 10 Bd Tonnellé, 37032 TOURS cedex, France., - Dr Annick TOUTAIN, MD, Service de Génétique, Centre Hospitalier Universitaire, 2 Bd Tonnellé, 37044 Tours cedex 9, France.

## Abstract

The ionotropic glutamate delta receptor GluD1, encoded by the *GRID1* gene, is involved in synapse formation, function, and plasticity. GluD1 does not bind glutamate, but instead cerebellin and D-serine, which allow the formation of trans-synaptic bridges, and trigger transmembrane signaling. Despite wide expression in the nervous system, pathogenic *GRID1* variants have not been characterized in humans so far. We report homozygous missense *GRID1* variants in five individuals from two unrelated consanguineous families presenting with intellectual disability and spastic paraplegia, without (p.Thr752Met) or with (p.Arg161His) diagnosis of glaucoma, a threefold phenotypic association whose genetic bases had not been elucidated previously. Molecular modeling indicated that Arg161His and Thr752Met mutations alter the hinge between GluD1 cerebellin and D-serine binding domains and the stiffness of this latter domain, respectively. Expression, trafficking, physical interaction with metabotropic glutamate receptor mGlu1, and cerebellin binding of GluD1 mutants were not conspicuously altered. Conversely, we found that both GluD1 mutants hampered signaling of metabotropic glutamate receptor mGlu1/5 via the ERK pathway in neurons of primary cortical culture. Moreover, both mutants impaired dendrite morphology and excitatory synapse density in neurons of primary hippocampal culture. These results show that the clinical phenotypes are distinct entities segregating in the families as an autosomal recessive trait, and caused by pathophysiological effects of GluD1 mutants involving metabotropic glutamate receptor signaling and neuronal connectivity. Our findings unravel the importance of the GluD1 receptor signaling in sensory, cognitive and motor functions of the human nervous system.

## Introduction

Intellectual disability (ID) and spastic paraplegia (SPG) are central nervous system disorders with marked clinical and genetic heterogeneity. In both groups, non-specific and syndromic forms have been described with numerous genes identified in the past few years (Ellison *et al*., 2013; Elsayed *et al*., 2021). The association of SPG with ID or MR (mental retardation, the out of use designation of ID) is frequent with 106 and 127 entries in the OMIM (Online Mendelian Inheritance in Man) database, respectively. Conversely, the triple combination of ID, SPG and glaucoma appears only once (OMIM#278050) with the description of two affected families: in four patients of both sexes in two sibships of a large inbred Swedish pedigree (Heijbel and Jagell, 1981), and in three male Canadian siblings born to first-cousin parents (Chenevix-Trench *et al*., 1986). Although the consanguinity and the presence of affected females in these families suggest an autosomal recessive inheritance, the genetic basis of this distinct entity is still unknown.

The glutamate delta receptors GluD1 (encoded by the *GRID1* gene) and GluD2 (*GRID2* gene) belong to the family of ionotropic glutamate receptors (iGluRs), which consist in homo- or heterotetrameric arrangements of subunits, and play key roles in synaptic transmission and plasticity (Traynelis *et al*., 2010; Yuzaki and Aricescu, 2017; Burada *et al*., 2021). GluDs do not bind glutamate but, instead, the binding of cerebellin and D-serine on distinct extracellular domains cooperatively gate GluD ion channels, whose opening is alternatively triggered by activation of Gq-coupled metabotropic glutamate receptors (mGlu1/5), or α1-adrenergic receptors (Ady *et al*., 2014; Benamer *et al*., 2018; Gantz *et al*., 2020; Carrillo *et al*., 2021). The binding of these ligands also triggers or modulates metabotropic signals, cerebellin additionally enabling postsynaptic GluDs to participate in excitatory synapse formation/stabilization via attachment with presynaptic neurexin (Yuzaki and Aricescu, 2017; Tao *et al*., 2018; Andrews and Dravid, 2021; Burada *et al*., 2021; Dai *et al*., 2021). GluD1 and GluD2 are widely expressed in the brain at excitatory postsynaptic sites, but GluD1 predominates over GluD2 outside the cerebellum, notably in the forebrain (Konno *et al*., 2014; Hepp *et al*., 2015; Nakamoto *et al*., 2019). However, truly pathogenic *GRID1* gene mutations have not been reported in human disease so far, in contrast with *GRID2* gene mutations (e.g. Hills *et al*., 2013; Utine *et al*., 2013; Maier *et al*., 2014; Coutelier *et al*., 2015; Grigorenko *et al*., 2022). Yet, the implication of *GRID1* in human disorders is suggested by Genome-Wide Association Studies showing that single-nucleotide polymorphisms and copy number variations in *GRID1* are risk factors for neuropsychiatric disorders (Fallin *et al*., 2005; Guo *et al*., 2007; Treutlein *et al*., 2009; Glessner *et al*., 2009; Cooper *et al*., 2011), and by the observation that *Grid1*^*−/−*^ mice exhibit abnormal behaviors, deficits in learning and memory, and alterations of dendritic spines, synapses, and mGlu1/5 signaling (Yadav *et al*., 2012; Yadav *et al*., 2013; Gupta *et al*., 2015; Suryavanshi *et al*., 2016; Liu *et al*., 2020; Andrews and Dravid, 2021)

Here, we report the identification of homozygous missense variants in the *GRID1* gene by genome-wide linkage analysis and/or whole exome sequencing (WES) in siblings from two unrelated consanguineous families presenting with mild or moderate ID, non- or slowly-progressive SPG, with (p.Arg161His) or without (p.Thr752Met) diagnosis of open angle glaucoma. Molecular modeling indicated that the mutations alter structural interactions within the extracellular domain of GluD1. Expression of GluD1 mutants in mouse primary neuronal cultures revealed that the mutations lead to impaired mGlu1/5 signaling, dendrite morphology, and excitatory synapse density.

## Materials and methods

### Patients

Written informed consent for genetic analysis was obtained from all participants or their legal guardians according to the Declaration of Helsinki and following Institutional Review Board (IRB)-approved protocols in the Centre Hospitalier Universitaire de Tours medical center (Family A) and the Hadassah Medical Center (Family B).

### Animals

Animal breeding and euthanasia were performed in accordance to European Communities Council Directive 86/609/062. *Grid1* KO mice (Gao *et al*., 2007; gift from Jian Zuo, Memphis, TE, USA) were genotyped as described (Hepp *et al*., 2015). Homozygous *Grid1* KO mouse embryos were obtained from breeding heterozygous parents. Wild-type (wt) mice were purchased from Janvier Labs. All mice had C57BL/6 background.

### Genome wide-linkage analysis and whole exome sequencing

Genomic DNA samples were extracted from peripheral blood following standard protocols. Genotyping of Family A (three affected children, one healthy child and both consanguineous parents) was performed on Genechip® human 250K NspI array (Affymetrix) according to the manufacturer’s instructions. Briefly 250 ng of genomic DNA were restricted with NspI. NspI adaptators were then ligated to restricted fragments followed by PCR using universal primer PCR002. PCR fragments were purified and 90 µg were used for fragmentation and end-labelling with biotin using Terminal Transferase. Labelled targets were then hybridized overnight to Genechip® human 250K NspI array (Affymetrix) at 49°C. Chips were washed on the fluidic station FS450 following specific protocols (Affymetrix) and scanned using the GCS3000 7G. The image was then analyzed with GCOS software to obtain raw data (CEL files). Genotypes were called by the Affymetrix GType software using Dynamic Model (DM) and Bayesian Robust Linear Model with Mahalanobis (BRLMM) mapping algorithms. Homozygosity regions were obtained using MERLIN software assuming a recessive model with complete penetrance (disease allele frequency of 0.0001).

WES study was performed using Agilent SureSelect Human All Exon kit (V2; Agilent technologies). Genomic DNA was captured with biotinylated oligonucleotides probes library (Agilent technologies), followed by paired-end 75 bases massive parallel sequencing on Illumina HiSEQ 2000. Image analysis and base calling were performed using the Illumina Real-Time Analysis Pipeline version 1.14 with default parameters. Sequencing data was analyzed according to the Illumina pipeline (CASAVA1.7) and aligned with the Human reference genome (hg19) using the ELANDv2 algorithm. Genetic variation annotation was performed with the IntegraGen in-house pipeline (IntegraGen). Filtering was performed using Eris software (IntegraGen) with an autosomal recessive hypothesis. Variants with minor allele frequency (MAF) >1% in either the 1000 Genomes Project, the EXAC, or the gnomAD databases were excluded. Genetic segregation of the candidate variant with the disease in Family A was confirmed by Sanger sequencing of *GRID1* exon 3.

For Family B, DNA sample of the proband was shipped to Otogenetics, USA (CLIA lab). ~50 Mb of genomic DNA were captured on HiSeq 2500. Fragments were read 100-125 bp, paired end. The sample was uploaded onto DNAnexus software and 71.5 million reads were aligned to the reference human genome (Hg19) (Mean on target coverage, X118). Variants which were low covered, off target (>6bp from splice site), synonymous, heterozygous, predicted as benign, MAF>0.5% on ExAC and MAF>4% in the Hadassah in-house dbSNP were removed. Thirty-one homozygous variants survived this filtering.

### Molecular modeling of GluD1^R161H^ mutant structure

The protein was generated using Rat GluD1 receptor in complex with 7-chloro-kynurenate and calcium ions (PDB codes: 6KSS and 6KSP) as structure templates (Burada *et al*., 2020). The system with proteins and ligands was prepared in the CHARMM-GUI web server (Jo *et al*., 2008) in order to generate a membrane around the protein and solvate with water and ions. A heterogeneous membrane made of POPC was chosen, and a TIP3 water model with NaCl (0.15 M) counter ions was chosen for the solvation. The system was typed with a CHARMM36m force field, and NAMD protocol was used. The system was equilibrated through six constrained simulations for a total of 500 ps by gradually diminishing the force constraints at each steps. The following constraints were applied (each value represents an equilibration step): protein backbone (5/2.5/1/0.5/0.1 kcal/mol), protein side chains (5/2.5/1.25/0.5/0.25/0.05 kcal/mol), lipid heads (5/5/2/1/0.2/0 kcal/mol), and dihedral bonds (500/200/100/100/50/0 kcal/mol). Then, a production dynamic of 5 ns was carried out in *NPT* conditions at 303.15 K without any constraints.

Mutant models were generated using Built Mutant protocol from Discovery Studio 2019. A set of 100 structures was created and ranked for their Dope score. The best model was then minimized using Adopted Basis Newton-Raphson algorithm (a Newton-Raphson algorithm applied to a subspace of the coordinate vector spanned by the displacement coordinates of the last positions) until a RMS gradient of 0.001 was obtained. Molecular docking experiments of D-Serine, glycine and kynurenic acid at the active site were performed as described (Ducassou *et al*., 2015, Dhers *et al*., 2017), using default parameters from CDocker (Wu *et al*., 2003) with Discovery Studio 2020 and a sphere radius of 10 Å in rigid mode. Flex Dock (Discovery Studio) was used for ligand-protein flexible docking.

### Plasmids and viruses

The following plasmids encoding mouse wild-type GluD1 (GluD1^WT^), mouse GluD1 variants, rat mGlu1a, or GFP under control of the cytomegalovirus (CMV) promoter were used for transfection of HEK293 cells: pcDNA3.1-HA-GluD1^WT^, pcDNA3.1-HA-GluD1^R161H^, pcDNA3.1-HA-GluD1^T752M^, pRK5-HA-mGlu1a-Venus; or of neuronal primary cultures: pmaxGFP (Lonza), pCMX-GFP (Umesono 1991; Drobac *et al*., 2010), pCMV-HA-GluD1^WT^, pCMV-HA-GluD1^R161H^; or of both HEK293 cells and neuronal primary cultures: pDEST26-GluD1^WT^, pDEST26-GluD1^R161H^, pDEST26-GluD1^T752M^. The hemagglutinin (HA) epitope YPYDVPDYA was inserted just after the predicted signal peptides of GluD1 and mGlu1a, this latter additionally comprising the Venus GFP variant fused to its C-terminus. Plasmids pRK5-HA-mGlu1a-Venus, pDEST26-GluD1^WT^, and pcDNA3.1-HA-GluD1^WT^ have been described previously (Perroy *et al*., 2008, Benamer *et al*., 2018). The R^161^H and T^752^M mutations were introduced in GluD1 through site-directed mutagenesis using the QuikChange II XL kit (Agilent Technologies). For generating pCMV-HA-GluD1^WT^, the full length coding sequence of the *Grid1* cDNA (Genbank accession number: BC167177) was PCR amplified from clone A230054J23 (Mus musculus adult male hypothalamus cDNA, RIKEN full-length enriched library, Refseq AK138279, Source BioScience) and inserted into the pCMV-HA-C plasmid (Clontech). A stop codon was then added at the end of the *Grid1* coding sequence upstream of the plasmidic HA tag, before inserting a HA tag after the predicted signal peptide. All constructs were verified with DNA sequencing.

Recombinant lentiviruses LV-PGK-GluD1^WT^-ires-GFP, LV-PGK-GluD1^R161H^-ires-GFP and LV-PGK-GluD1^T752M^-ires-GFP were used for transduction of neurons in primary cortical cell culture. These lentivectors were generated exactly as described (Benamer *et al*., 2018) for co-expression of GluD1^WT^/GluD1^R161H^/GluD1^T752M^ and GFP driven by the PGK promoter. Recombinant lenti pseudo-virions were produced at the Viral Vector and Gene Transfer facility of the Necker Institute (IFR94, Paris, France).

### HEK293T cell culture and transfection

HEK293T cells (ATCC Number: CRL-3216, authenticated using Short Tandem Repeat analysis by the ATCC cell authentication service, mycoplasma-free) were cultured in Dulbecco’s modified Eagle’s medium (DMEM) supplemented with 10% fetal bovine serum, 100 U/ml penicillin and 100 µg/ml streptomycin (Life Technologies). For immunostaining or cerebellin binding experiments, cells were seeded at 8.10^5^ cells per well on glass coverslips coated with poly D-lysine (Sigma Aldrich P7280) and cultured in 12-well plates. For membrane protein isolation and immunoprecipitation experiments, cells were seeded in 10 cm dishes coated with poly D-lysine at a density of 2.10^6^ cells/dish. Transient plasmid transfection was performed the next day using the calcium phosphate precipitation method (6 µg plasmid per 12 well-plate or per dish) or using Lipofectamine 2000 (Invitrogen, 2,5 µg plasmid and 6µL reagent per 6 well-plate). Plasmids encoding mGlu1-YFP and GluD1 were mixed at a ratio 1:1 for co-transfection. Culture medium was renewed 6 h after transfection, and cells cultured overnight.

### Immunostaining on HEK cells

Transfected HEK cells were fixed with 4% paraformaldehyde in 0.1 M sodium phosphate buffer (PB) during 20 min, and then washed with Dulbecco’s phosphate-buffered saline (D-PBS). All the procedure was performed at room temperature. After fixation, cells were incubated in PBS containing fish skin gelatin (2g/l) and Triton X100 0,25% (PBS-GT) for 1 hour. Triton X100 was omitted from incubation medium (PBS-G) when cells were not permeabilized. Next, cells were incubated for 2 to 4 hours with primary antibodies (see **Table 1** for antibodies) diluted in PBS-GT/PBS-G, washed 3 times 15 minutes with PBS, and incubated with secondary antibodies (**Table 1**) and DAPI nuclear stain (300 nM, Invitrogen) diluted in PBS-GT for 2 hours. After PBS washes, samples were mounted on glass slides using Fluoromount-G (Biovalley 0100-01), and images were acquired using an epifluorescence microscope (DMR, Leica), or a confocal microscope (SP5, Leica).

**Table 1:** Antibodies.

| **Primary Antibodies** | **reference** | **Immuno-precipitation** | **Immuno-fluorescence** | **Western blot** |
| --- | --- | --- | --- | --- |
| *rabbit anti-GluD1* | *Hepp et al. 2015* | *7 µg/250µg prot* | *1/10,000* | *1/10,000* |
| *rabbit Anti-Mouse IgG* | *Jackson Immunoresearch 315-005-003* | *2µg/2µg mouse anti-HA* |  |  |
| *mouse anti-HA* | *Biolegend clone 16B12* | *2 µg/250µg prot* | *1/2000* | *1/5000* |
| *rabbit anti-HA* | *Clontech Takara Bio 631207* |  | *1/500* | *1/1000* |
| *Chicken anti-GFP* | *Aves GFP-1020* |  | *1/1000* |  |
| *Rabbit anti-GFP* | *Chromtek PABG1* |  | *1/1000* |  |
| *Mouse anti MAP2* | *Sigma M9942* |  | *1/1000* |  |
| *Rabbit anti phosphoERK1/2* | *Cell Signaling 4370S* |  |  | *1/2000* |
| *Mouse anti ERK1/2* | *Cell Signaling 4696S* |  |  | *1/2000* |
| *Mouse anti-beta-actin HRP-conjugated* | *Sigma-Aldrich A3854* |  |  | *1/50000* |
| **Secondary Antibodies** |  |  |  |  |
| *Goat anti-rabbit HRP-conjugated* | *Promega W4011* |  |  | *1/2500* |
| *Goat anti-Mouse IgG DyLight 680* | *Thermo Fisher 35519* |  |  | *1/5000* |
| *Goat anti-Rabbit IgG DyLight 800* | *Thermo Fisher SA5-10036* |  |  | *1/5000* |
| *Donkey anti-rabbit Alexa Fluor 488* | *Thermo Fisher A21206* |  | *1/2000* |  |
| *Goat anti-Rabbit IgG Alexa Fluor 555* | *Thermo Fisher A21430* |  | *1/2000* |  |
| *Donkey anti-rabbit Alexa Fluor 594* | *Interchim FP-SD5115* |  | *1/500* |  |
| *Goat anti-Mouse IgG Alexa Fluor 488* | *Thermo Fisher A11029* |  | *1/2000* |  |
| *Goat anti-Mouse IgG Alexa Fluor 555* | *Thermo Fisher A21422* |  | *1/2000* |  |
| *Goat anti-Mouse IgG Alexa Fluor 647* | *Thermo Fisher A21235* |  | *1/2000* |  |
| *Goat anti-Chicken IgY Alexa Fluor 488* | *Thermo Fisher A11039* |  | *1/2000* |  |

### Cerebellin binding on HEK cells

HEK cells expressing GluD1 or GluD1^R161H^ were incubated for 1 hour at 35 °C in culture medium containing 20 µg/ml recombinant human HA-tagged Cerebellin 1 (Cbln1, Biotechne 6934-CB-025). After two washes with ice-cold PBS, cells were fixed and processed for immunostaining as described above using rabbit anti-GluD1 and mouse anti-HA primary antibodies (**Table 1**).

### Isolation of membrane proteins from HEK cells and western blotting

Total membrane proteins were extracted from HEK cells expressing HA-GluD1^WT^, HA-GluD1^R161H^ or HA-GluD1^T752M^ using the MEM-Per™ Plus Membrane Protein extraction kit (Thermoscientific) according to manufacturer’s protocol. Proteins lysates were separated on 4-20% Mini-PROTEAN**®** TGX Stain-Free Precast electrophoresis Gels (Bio-Rad) and transferred using Trans Blot Turbo system (Bio-Rad) on nitrocellulose membranes (Bio-Rad). Membranes were then incubated in blocking buffer with 5% milk in a mixture of Tris-buffered saline and Tween 0,002% (TBST, Fisher) for 1 hour at room temperature. Next, membranes were incubated with rat anti-HA antibody (**Table 1**) overnight at 4°C in 5% milk diluted in TBST. After three washes of 10 min, membranes were incubated in 5% milk-TBST with secondary anti-rabbit and anti-beta-actin antibodies conjugated with horseradish peroxydase (HRP, **Table 1**) for 45 min. HRP was revealed through chemiluminescence using Clarity™ Western ECL substrate (Bio-rad), visualized on a ChemiDoc™ Touch imaging system (Bio-rad), and quantified using the ImageJ software (U.S. National Institutes of Health, Bethesda, MD, USA; http://rsbweb.nih.gov/ij/).

### Immunoprecipitation from HEK cells

HEK cells co-expressing HA-mGlu1a-Venus and GluD1, GluD1^R161H^, or GluD1^T752M^ were washed twice with ice cold PBS and lysed in 500 µl lysis buffer containing 50mM Tris-HCl pH 7.5, 150mM NaCl, 1% Nonidet P40, 0.5% sodium deoxycholate, and protease inhibitor (Complete Ultra Tablets, Roche) according to manufacturer’s instructions. The whole immunoprecipitation procedure was carried out at 4 °C. Lysates were centrifuged 13000g for 15 min and protein concentration was determined in the supernatant by the Bradford’s method using BSA as standard. Supernatants were then pre-cleared with Protein A Plus Agarose beads (Pierce). Specific immunoprecipitation were performed overnight by incubating 250 µg proteins of the precleared lysates with specific antibodies or control rabbit anti-mouse antibodies (**Table 1**). Protein complexes bound to rabbit anti-GluD1 antibodies were precipitated with Protein A Plus agarose beads for 4 h. Protein complexes bound to mouse anti-HA antibodies were precipitated with beads coupled to rabbit anti-mouse antibodies. Precipitates were washed twice with lysis buffer, twice with 50 mM Tris Hcl pH7.5, 500 mM Nacl, 0.1% Nonidet P40, 0.05% Sodium deoxycholate and once with 50 mM Tris Hcl 0.1% Nonidet P40, 0.05% Sodium deoxycholate. Proteins were eluted from the beads with 30 µl LDS sample buffer (Invitrogen), separated on 4-15 % polyacrylamide gels (Biorad), and transferred onto nitrocellulose membranes. Western blots were carried out using standard protocols and antibodies listed in **Table 1**. Detection was performed with the Odyssey detection system (LI-COR Bioscience) using secondary anti-IgG antibodies coupled to infrared dyes (**Table 1**). Band intensity was determined using ImageJ.

### Primary cortical or hippocampal cell cultures

All components for cell cultures were from Thermo Fisher Scientific unless otherwise stated. Primary cortical or hippocampal cell cultures were prepared essentially as described (Ung *et al*., 2018) from E17-E18 *Grid1*^−/−^ or *Grid1*^+/+^ mice embryos, respectively. Cortices and hippocampi were dissected in ice cold PBS containing 100 U/ml penicillin and 100 µg/ml streptomycin, and kept in Hibernate E medium supplemented with 2% B27, while genotyping using the Phire Animal Tissue Direct PCR Kit (Thermo Fisher, for primers see Hepp *et al*., 2015). Tissues were pooled, dissociated with papain (Worthington). Tissues were then triturated in DMEM-F12 containing 10 % heat inactivated fetal calf serum and cells transferred to a new tube and centrifuged 250 g for 4 minutes. Cells were resuspended in Neurobasal medium supplemented with 2% B27, 0.5 mM glutamax (complete Neurobasal medium), and counted. Cortical cells were plated at 10^6^ cells per dish on 35 mm culture dishes coated with poly D-lysine and laminin (Sigma Aldrich). Hippocampal cells were seeded at a density of 6×10^4^ cells/500 µl medium per well on glass coverslips coated with poly D-lysine and laminin in 24 well plates. Cells were grown at 35°C, under 5% CO2 atmosphere. Half of the medium was changed every 3 to 4 days.

### Viral transduction of cortical cell cultures and test of mGlu1/5 signaling

At 10 days *in vitro* (DIV), cultures were transduced with LV-PGK-GluD1-ires-GFP, LV-PGK-GluD1^R161H^-ires-GFP, or LV-PGK-GluD1^T752M^-ires-GFP recombinant pseudo-virions at a density of infection of 1:1, and then cultured for 4 additional days.

For measurements of lentiviral transduction efficiency, cultures were next fixed and processed for DAPI staining and immunolabelling (see **Table 1**) as described above for HEK cells.

For test of mGlu1/5 signaling, cultures were next rinsed once with warm HBSS-TTX-APV medium containing 2 mM Ca2+, 1 mM Mg2+, 300 nM TTX (Latoxan), and 50 µM of the NMDAR antagonist APV (Hello Bio). Cells were then incubated for 1 hour at 35° in HBSS-TTX-APV. The medium was next removed, and cells were incubated for 5 minutes at 35°C in HBSS-TTX-APV medium, in the presence or absence of RS-3,5-dihydroxyphenylglycine 2 (DHPG, 100 µM, Hello Bio). Cells were then lysed in ice cold 50 mM Tris-HCl pH 7.5, 150 mM NaCl, 1% Nonidet P40, 0.5% sodium deoxycholate, supplemented with protease (Complete Ultra Tablets, Roche) and phosphatase inhibitors (PhoStop, Roche) according to manufacturer’s instructions. Protein concentration was determined using the Bradford’s method with BSA as standard. Proteins (10 µg/lane) were separated by SDS-PAGE, transferred onto nitrocellulose sheets, and western blots were carried out using standard protocols. Primary antibodies and secondary anti-IgG antibodies coupled to infrared dyes are listed in **Table 1**. Detection was performed with the LI-COR Odyssey detection system. Band intensity was determined using the ImageJ software.

### Plasmid transfection of hippocampal cell cultures and analyses of neurites and excitatory synapses

Cultures were transfected at DIV4 for morphometric analyses of dendrites, and at DIV11-DIV13 for analyses of spine morphology and synapse counting. Cells were transfected with plasmid pCMX-GFP alone or in combination with pDEST26-GluD1^WT^, pDEST26-GluD1^R161H^, or pDEST26-GluD1^T752M^ (1:1 ratio) using Lipofectamine 2000 (Invitrogen). Plasmids pmaxGFP, pCMV-HA-GluD1^WT^ and pCMV-HA-GluD1^R161H^ were used instead of above plasmids for experiments shown in **Figures S3 and S8**. Lipofectamine (1 µl/well) and plasmids (500 ng/well) were diluted in Neurobasal medium (100 µl/well). Prior to transfection, 300 µl medium was collected from each well and diluted by half with fresh complete Neurobasal medium. This conditioned medium was kept in the incubator for the duration of the transfection. Next, 200 µl complement-free Neurobasal medium and 100 µl of the lipofectamine-DNA solution was added in each well. After 2 h incubation at 35°C, cells were washed twice with complete Neurobasal medium before adding 500 µl conditioned medium. Cells were then incubated for 48 h before fixation. Cells were fixed with 4 % paraformaldehyde for 20-30 min, permeabilized with PBS-GT unless otherwise stated, and processed for immunolabelling and DAPI staining as described above for HEK cells. Immunolabelling of co-transfected cultures using chicken anti GFP, and rabbit anti-GluD1 or rat anti-HA primary antibodies (**Table 1**), demonstrated that more than 96 % of GFP-expressing neurons also over-expressed either GluD1^WT^, GluD1^R161H^, GluD1^T752M^, HA-GluD1^WT^, HA-GluD1^R161H^, or HA-GluD1^T752M^.

***For morphometric analyses of dendrites***, images of isolated GFP-expressing neurons were acquired with an epifluorescence microscope (DMR, Leica). The Sholl analysis was performed upon conversion to binary images using the SNT module of ImageJ/Fiji software (https://imagej.nih.gov/ij/, Schindelin *et al*., 2012; Schneider *et al*., 2012; Ferreira *et al*., 2014). An ROI delimiting the soma was used to define the cell center from which concentric circles of 20 pixels (5 µm) apart were drawn on a radius of 700 pixels. For each cell, the number of neurites crossing along the radius and the total crossings were determined. The total length of neurites per cell was manually determined using the segmented line tool of ImageJ/Fiji. Lines were converted to ROIs to determine the length of all the segment per cells in order to sum them up. Neurites extending beyond the field were not included.

***For analyses of dendritic spine morphology***, images of isolated GFP-expressing spiny dendrites were acquired by confocal microscopy (TCS SP8-STED, Leica) using a 63X objective with a zoom of 2 and were z-sectioned at 0.3 µm increments. Morphological analysis of the GFP-labelled spines was performed manually according to Zagrebelsky *et al*. (2005), based on measurements of spine length and of the ratio between neck and head diameters of the spine. We distinguished immature spines comprising both long thin (length: 1<x<3 µm, head/neck diameter<2) and filopodia-shaped (length >3 µm) spines, versus mature spines comprising both mushroom-shaped (length: 1<x<3 µm, head/neck diameter>2) and stubby (length<1 µm) spines.

***For synapse counting***, cells were labelled using primary antibodies: chicken anti GFP, mouse anti Bassoon, rabbit anti Homer1, and secondary antibodies: goat anti-chicken Alexa 488, goat anti-mouse-RRX, goat anti-rabbit-Alexa 647 (**Table 1**). Images were acquired with a DMR Leica epifluorescence microscope. The density of glutamatergic synapses was measured by counting manually Homer1/Bassoon co-labelled spots present on merged images of GFP positive dendrites processed with ImageJ/Fiji. Only spiny neurons exhibiting a pyramidal cell-like morphology with pyramidal-shaped soma and prominent apical dendrite were analysed.

### Statistical analyses

All experiments were repeated at least three times, and GraphPad prism6 software (Instat) was used for statistical analyses and graphical representations. When d’Agostino-Pearson normality tests were successfully passed, we conducted parametric test using One-way ANOVA. Then, Tukey’s post hoc method was used to determine statistical significance in multiple comparisons and to reveal the contribution of the genotype in the variability between each test. For samples that did not pass the normality test, we used Kruskal-Wallis method followed by Dunn’s post hoc test. Results are given as mean ± standard error of the mean. Differences were considered significant if p<0.05.

## Results

### Clinical description of the families

Family A included three affected siblings born to consanguineous parents. The affected siblings presented with non- or slowly-progressive SPG diagnosed in infancy with no other neurological signs, mild/moderate ID with normal occipitofrontal circumference, and juvenile open angle glaucoma causing severe visual impairment. Overall, the clinical pictures of the siblings were strikingly similar to earlier descriptions of this syndrome (Heijbel and Jagell, 1981; Chenevix-Trench *et al*., 1986). Brain MRI, electromyography, metabolic investigations, and standard chromosome analysis of the three siblings were normal. Linkage to genes *ARX, XNP, PLP* and *L1CAM* was excluded, sequencing of *MECP2* did not detect a causative variant, and high-resolution array-Comparative Genomic Hybridization (CGH) analysis (Agilent CGH array 1M) did not reveal any pathogenic copy number variation related to the disease.

Family B included two affected siblings born to consanguineous parents. The proband presented with global developmental delay, spastic paraplegia, dysmorphic features, and minor skeletal anomalies. The other sibling was similarly affected. Ophthalmologic examination could not be performed on either sibling. Initial genetic investigations for the proband of Family B included chromosomal karyotype analysis which was normal, as well as CGH analysis, which was considered normal but notable for an intronic 50Kb deletion in 7q36.2 encompassing the *DPP6* gene (arr:7q36.2(153,921,762-153,951,944)X1).

The clinical features for the five affected individuals are summarized in **Table 2**, together with their side-by-side comparison with observations by Heijbel and Jagell (1981) and Chenevix-Trench *et al*. (1986). Additional information on family pedigrees and on medical conditions is available upon request to corresponding authors.

**Table 2:**
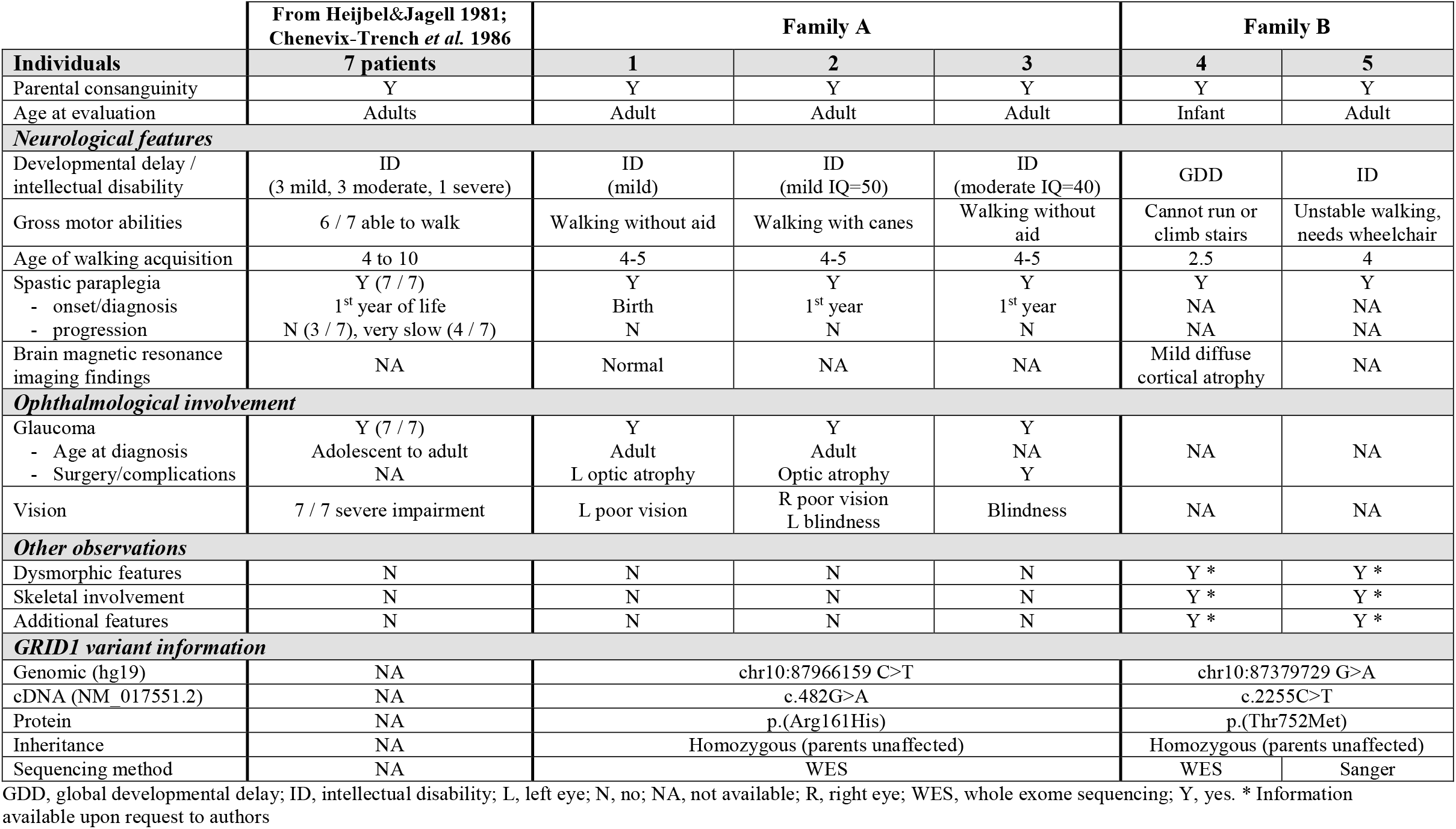
Clinical and genetic features of patients with ID and SPG with or without glaucoma, from present and earlier reports.

### Identification of homozygous variants in *GRID1*

As the consanguinity in both families suggested an autosomal recessive inheritance transmission model, we performed genome-wide single nucleotide polymorphism (SNP) genotyping for Family A in the three affected siblings, in one healthy sibling and in both parents. Two homozygous regions with significant linkage were found at chromosomes 10 and 12. The first region of 30.6 Mb was located at 10q23.1-q25.2 (hg19, chr10:81864184-112544655) and delimited by rs11201697 and rs7077757 markers. This region encompasses around 250 genes and miRNAs, and provides a peak multipoint Logarithm of the ODds (LOD) score of 2.53 in the family (**Figure S1**). The second homozygous genetic interval spans 161 kb at chromosome 12q24.33 (from hg19, chr12:130120222 to chr12:130281773) between rs10773690 and rs4759984 markers, and contains the second exon of the *TMEME132D* gene (**Figure S1**). We next performed a WES analysis on two affected siblings in Family A and one of their parents. Variants were filtered according to quality criteria, potential pathogenicity, and population frequency (Minor Allele Frequency <1%). This allowed the identification of a homozygous missense mutation of the *GRID1* gene (NM_017551.2: c.482G>A, p.Arg161His; hg19, chr10:87966159 C>T) within the 10q22q23 candidate region (**Figure 1A**). This variant segregated in an autosomal recessive manner in all affected members of the family (**Table 2**), with both parents and an unaffected sibling found to be heterozygous carriers, and is predicted as “Disease causing” by *Mutation Taster* (score 0.9697). Referred in dbSNP (rs771100097), the p.Arg161His *GRID1* variant is found at heterozygous state in 4 individuals in gnomAD database (v2.1.1, Minor Allele Frequency, MAF=1.60e-5), but is not reported as homozygous.

**Figure 1:**
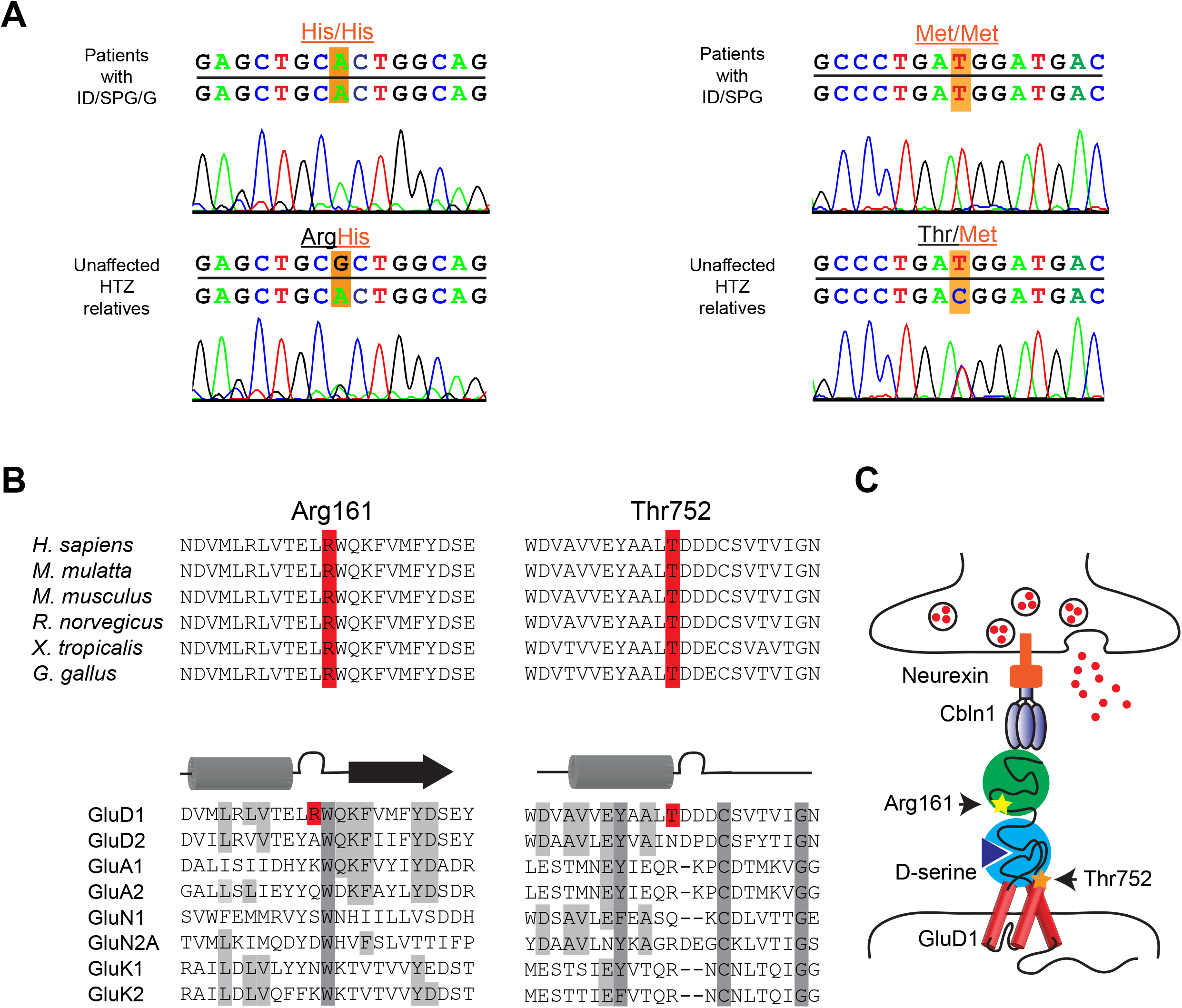
Homozygous *GRID1* variants p.Arg161His and p.Thr752Met causing ID and SPG with or without Glaucoma. **(A)** Sanger sequencing electrophoregrams showing the *GRID1* homozygous missense mutations c.482G>A, p.Arg161His and c.2255C>T, p.Thr752Met in the affected patients and the heterozygous mutations in unaffected relatives. **(B)** Amino-acid alignments showing conservation of GluD1 R^161^ and T^752^ residues across species, but not among iGluR family members. **(C)** Spatial organization of the transsynaptic complex GluD1-cerebellin (Cbln1)-neurexin at the glutamatergic synapse. Note that R^161^ and T^752^ residues belong to cerebellin-binding (ATD) and D-serine-binding domains (LBD), respectively.

For Family B, single (proband-only) WES was pursued, and brought to the identification of a homozygous missense variant in *GRID1* (NM_017551.3: c.2255C>T, p.Thr752Met; hg19, chr10:87379729G>A) (**Figure 1A**). Using Sanger sequencing, this variant was confirmed to segregate with the disease in the family, with both affected siblings homozygous for the variant (**Table 2**), both parents and an unaffected sibling found to be heterozygous carriers, and three additional unaffected siblings wild type for the variant. The p.Thr752Met variant is only found at heterozygous state, in 11 individuals from gnomAD database (MAF=3.89e-5).

Finally, we sequenced a cohort of more than 200 patients affected with SPG, isolated or associated with ID, but this search failed to identify additional variants in *GRID1*.

### Structural impact of Arg161His and Thr752Met mutations on GluD1 extracellular domains

The p.Arg161His (R^161^H) and p.Thr752Met (T^752^M) mutations concern GluD1 amino acid residues conserved among vertebrate species, but not among iGluR subunits (**Figure 1B**), consistent with functional heterogeneity within this receptor family (Schmid and Hollmann, 2008; Traynelis *et al*., 2010; Burada *et al*., 2020). Based on GluD1 sequence and cryo-EM 3D structure (Burada *et al*., 2020; Burada *et al*., 2021), we assigned the R^161^ and T^752^ residues to the extracellular Amino Terminal Domain (ATD) and Ligand Binding Domain (LBD) of GluD1, which bind cerebellin and D-serine, respectively (**Figure 1C**). The R^161^ residue is situated at the interface between ATD and LBD, distant from ATD residues involved in cerebellin binding, whereas the T^752^ residue lies within the LBD (**Figure 2A, 2C**). We thus modelled the complete structure by generation of unresolved 3D loops crucial for GluD1 activation. In this full-length model, we characterized the possible impact of the mutations on the ATD, LBD, and their coordination, based on the 3D structure of GluD1 (Burada *et al*., 2020, see **Figure S2**). The modeling results indicate that the R^161^H mutation impacts interactions of the hinge between the two domains by modifying the binding pattern with the Q^416^, D^417^, and P^419^ residues of the loop linking ATD to LBD (**Figure 2B**), with possible consequences on the structural and functional cooperativity between the two domains (Elegheert *et al*., 2016; Burada *et al*., 2020; Carrillo *et al*., 2022). The T^752^M mutation results in additional interactions between lateral chains of M^752^, Y^748^ (in the same α helix) and I^729^ (in adjacent α helix) that could lead to a stiffening of the structure and thus a decrease in the flexibility of the peptidic backbone (**Figure 2D**). The molecular docking results also indicate that D-serine affinity for the LBD is decreased in GluD1^T752M^, but only slightly modified in GluD1^R161H^ (binding energy: wild-type GluD1 (GluD1^WT^), −107.9; GluD1^R161H^, −102.7; GluD1^T752M^, −68.8 kJ/mole), whereas binding of endogenous ligand glycine and of synthetic ligand 7-chloro-kynurenate to the LBD (Kristensen *et al*., 2016) are weakened by both mutations (from −100.7 and −108.8 kJ/mole in GluD1^WT^, to −76.8 and −79.1 kJ/mole in GluD1^R161H^, and to −65,7 and −77.8 kJ/mole in GluD1^T752M^, respectively). These results suggest that the R^161^H and T^752^M mutations can both affect GluD1 function by altering the transduction of ligand binding to transmembrane/intracellular signaling of this receptor.

**Figure 2:**
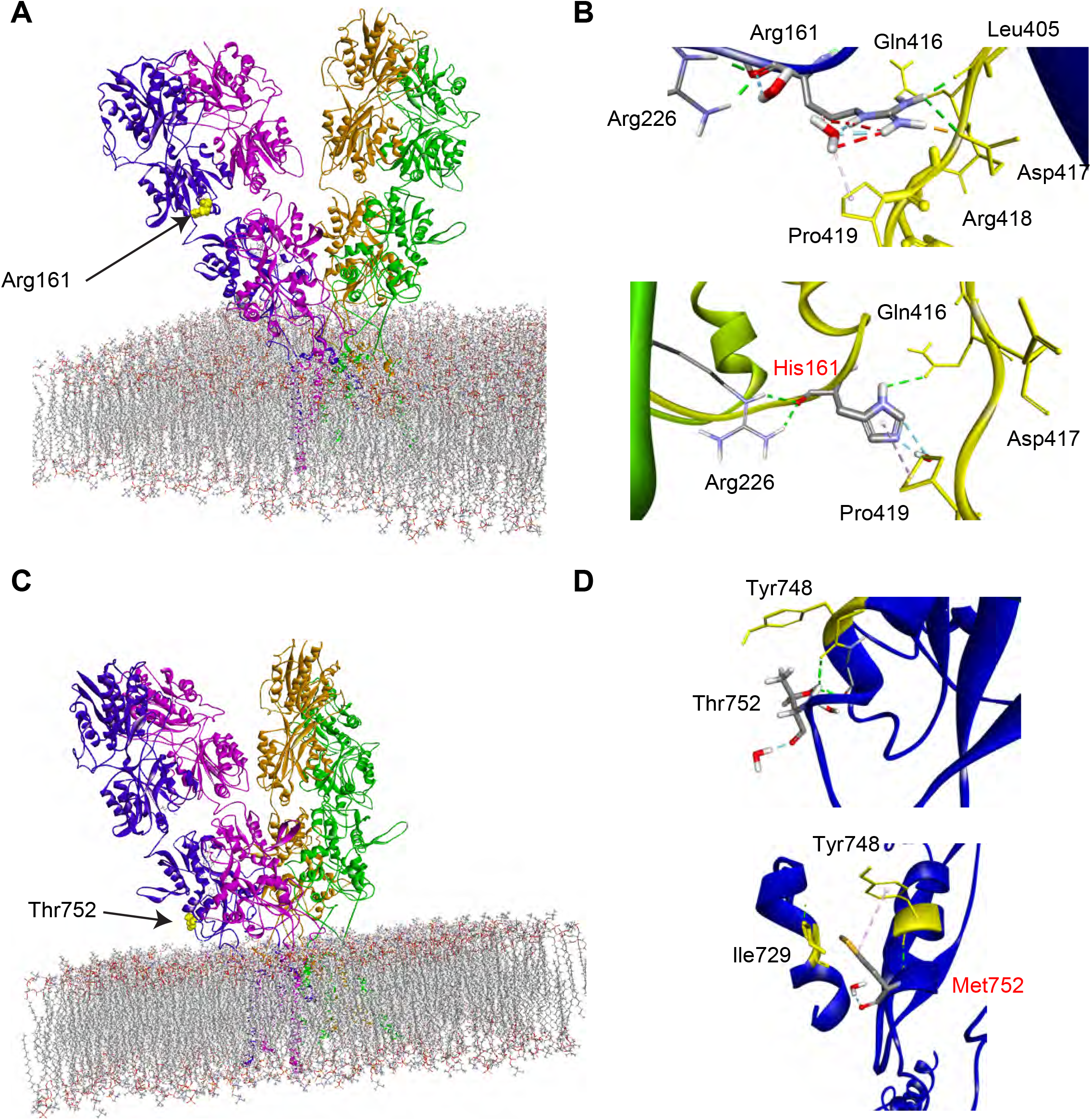
Modelling the structural impact of GluD1 R^161^H and T^752^M mutations on cerebellin-binding and D-serine-binding domains. **(A)** Structure of the GluD1 homotetramer sitting above the plasma membrane - adapted from Burada *et al*. (2020). Mutations affect residues situated at the interface (R^161^) between ATD and LBD extracellular domains, or within LBD (T^752^). **(B)** Predicted interactions of wt R^161^ and T^752^ residues, and of mutant H^161^ and M^752^ residues. Note that the R^161^H mutation suppresses interaction with D^417^ residue of the loop linking ATD to LBD, thereby changing loop conformation, whereas the T^752^M mutation results in supplementary interaction with I^729^ and Y^748^ residues of the LBD, thereby rigidifying this latter domain **(C and D)**.

### The GluD1 R^161^H and T^752^M mutations do not hamper cerebellin binding to GluD1

To gain further insight into the impact of the mutations on GluD1 function, we first compared the expression level and subcellular localization of GluD1^WT^, GluD1^R161H^ and GluD1^T752M^, bearing an N-terminal extracellular HA-tag, and expressed in HEK cells through plasmid transfection (see Methods). Western blot analyses indicated that HA-GluD1^WT^, HA-GluD1^R161H^, and GluD1^R161H^ did not conspicuously differ in amount, molecular weight, or insertion in cell membranes (**Figure S3A**), suggesting that GluD1 expression, stability, and membrane targeting is not affected by the R^161^H and T^752^M mutations. This latter point was further investigated using transient overexpression of HA-GluD1^WT^, HA-GluD1^R161H^, and HA-GluD1^T752M^ in mature hippocampal primary neuronal cultures from *Grid1*^+/+^ mice (see Methods). Anti-HA staining of non-permeabilized, putative excitatory neurons, revealed that HA-GluD1^WT^, HA-GluD1^R161H^, and HA-GluD1^T752M^ were all expressed at the neuronal plasma membrane and similarly distributed along dendritic shafts and spines (**Figure S3B**). These results suggest that the pathogenic effects of GluD1^R161H^ and GluD1^T752M^ mutant receptors do not result from deficits in their expression, stability or trafficking.

We next compared the ability of GluD1^WT^, GluD1^R161H^ and GluD1^T752M^ to bind cerebellins, through which postsynaptic GluDs anchor trans-synaptic scaffolds via attachment with presynaptic neurexin (Yuzaki and Aricescu, 2017; Tao *et al*., 2018; Dai *et al*., 2021). Cerebellin binding was tested by incubating HEK cells expressing GluD1^WT^, GluD1^R161H^ or GluD1^T752M^ through plasmid transfection, with recombinant HA-tagged Cerebellin 1 (Cbln1, see Methods). We found that both GluD1 mutants retained the cerebellin-binding capability of GluD1^WT^ as judged from similar anti-HA immunostaining of HEK cell membranes in the three conditions (**Figure S4**). These results indicate that cerebellin binding, and thus trans-synaptic scaffolding ability, is essentially preserved in GluD1 mutants, consistent with R^161^H and T^752^M mutations being distant from cerebellin-binding residues in the structure of the receptor.

### The GluD1 R^161^H and T^752^M mutations impair the modulation of mGlu1/5 signaling by GluD1

Both the binding of cerebellin and D-serine to GluDs trigger postsynaptic signaling relevant to synapse formation, stabilization, function and plasticity (Yuzaki and Aricescu, 2017; Tao *et al*., 2018; Dai *et al*., 2021; Burada *et al*., 2021; Carrillo *et al*., 2022). Our above molecular modeling results suggest that both mutations can affect transduction of ligand binding to GluD1 signaling. Because GluD1 associates both physically and functionally with mGlu1/5 receptors (Suryavanshi *et al*., 2016, Benamer *et al*., 2018), we tested the impact of GluD1 mutations on mGlu1/5 signaling via the ERK pathway, which is involved in the control of synapse formation and plasticity and is altered in ID (Impey *et al*., 1999; Sweatt, 2001; Davis and Laroche, 2006; Stoppel *et al*., 2017; Wilkerson *et al*., 2018; Lavoie *et al*., 2020).

We first verified that GluD1 mutants associate with HA-tagged mGlu1 upon co-expression in HEK cells (see Methods and **Figure S5A**). Using anti-HA or anti-GluD1 antibodies, we found that mGlu1 co-immunoprecipitated with GluD1^WT^, GluD1^R161H^, or GluD1^T752M^ with a similar efficiency (**Figure S5B**), indicating that the mGlu1-GluD1 physical interaction is not impaired by the GluD1 R^161^H and T^752^M mutations.

Next, GluD1^WT^, GluD1^R161H^, or GluD1^T752M^ were co-expressed with Green Fluorescent Protein (GFP) through viral transduction in primary cultures of cortical cells from *Grid1*^−/−^ mice (see Methods), in order to avoid influence of endogenous GluD1^WT^ on mGlu1/5 signaling in non-transduced cells. All GFP-labelled transduced cells examined were GluD1-immunopositive (**Figure 3A**), and the vast majority of neurons in these cultures were transduced (**Figure S6**). Following 5 min incubation in the presence/absence of the mGlu1/5 agonist DHPG (100 µM), cultures were processed for western blot and immunoquantification of the phosphoERK/ERK ratio (see Methods and example in **Figure 3B**). In GluD1^WT^- expressing cultures, DHPG treatment stimulated ERK signaling, as revealed by the strong increase of the phosphoERK/ERK ratio relative to mock-treated control cultures (DHPG: 221 ± 8 % of control; DHPG>control, p<0.05; n=27 control, n=27 DHPG-treated cultures, **Figure 3C**). The same paradigm elicited a significantly weaker stimulation of the ERK pathway in GluD1^R161H^- or GluD1^T752M^-expressing cultures (**Figure 3C**). Indeed, the increase of phosphoERK/ERK ratio by DHPG was only 161 ± 9 % of control in GluD1^R161H^-expressing cultures (DHPG-GluD1^WT^>DHPG-GluD1^R161H^, p<0.05; n=27 DHPG-GluD1^WT^, n=14 DHPG-GluD1^R161H^), and 180 ± 12 % of control in GluD1^T752M^-expressing cultures (DHPG-GluD1^WT^>DHPG-GluD1^T752M^, p<0.05; n=27 DHPG-GluD1^WT^, n=10 DHPG-GluD1^T752M^). These results indicate that the modulation by GluD1 of mGlu1/5 signaling via the ERK pathway is impaired by the GluD1 R^161^H and T752M mutations.

**Figure 3:**
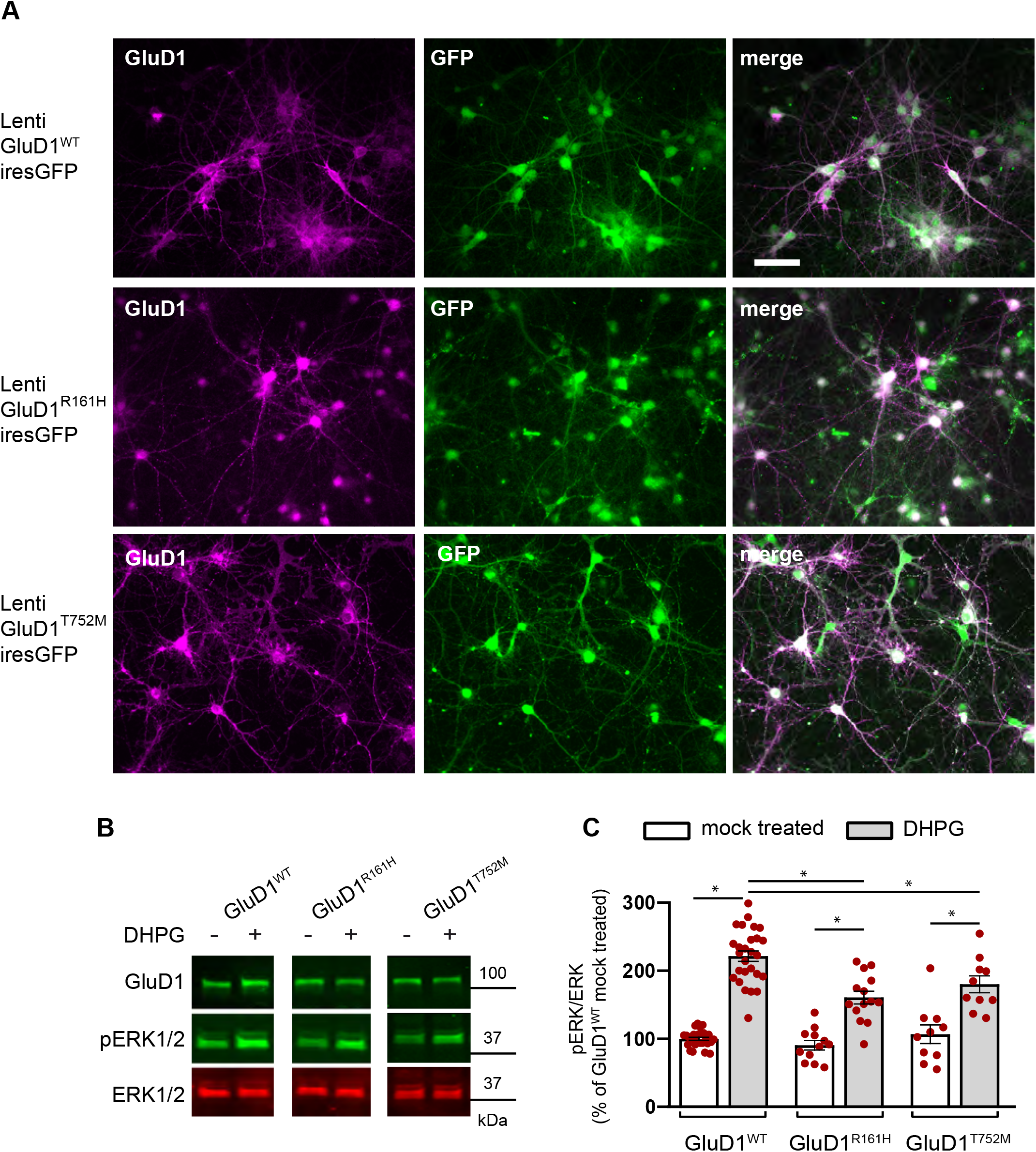
The R^161^H and T^752^M mutations hamper the modulation of mGlu1/5 signaling by GluD1. **(A)** Fluorescence pictures of primary cortical cell cultures from *Grid1*^−/−^ mouse co-expressing GluD1^WT^/GluD1^R161H^/GluD1^T752M^ and GFP following lentiviral transfer. **(B)** Western blot analysis of virally transduced cortical cultures following incubation in presence of the NMDAR antagonist APV (50 µM), with or without mGlu1/5 agonist DHPG (100 µM). Note the higher intensity of phosphoERK (pERK1/2), indicative of ERK signaling activation, following incubation with DHPG. **(C)** Summary of results obtained in mock-treated or DHPG-treated primary cortical cell cultures expressing GluD1^WT^ (n=27 and 27, respectively), GluD1^R161H^ (n=12 and 14, respectively), or GluD1^T752M^ (n=10 and 10, respectively). Note that the increase of the pERK/ERK ratio by DHPG was significantly larger in GluD1^WT^-expressing than in GluD1^R161H^-, or GluD1^T752M^-expressing cultures.

### The GluD1 R^161^H and T^752^M mutations impair dendrite morphology and excitatory synapse density

Alterations of dendritic spines and synapses are found in various forms of ID in humans and in ID mouse models (Banerjee *et al*., 2019; Bagni and Zukin, 2019; Lima Caldeira *et al*., 2019). We thus examined the impact of the GluD1 R^161^H and T^752^M mutations on neuronal dendrites and excitatory synapses using co-transfection of plasmids encoding GluD1^WT^, GluD1^R161H^, or GluD1^T752M^ together with a GFP-expressing plasmid in mature hippocampal primary neuronal cultures from wt mice (see Methods). We found that the vast majority of GFP-expressing neurons also over-expressed either GluD1^WT^ (96.8 ± 4.2 %, n=252), GluD1^R161H^ (96.3 ± 3.9 %, n=246), or GluD1^T752M^ (96.1 ± 4.7 %, n=250), as shown by dual GFP and GluD1 immunolabelling. Moreover, plasmid-driven expression of GluD1^WT^ and GluD1 mutants was largely superior to that of endogenous GluD1 (**Figure S7**), allowing the effect of recessive GluD1 R^161^H and T^752^M mutations to be evaluated in transfected wt neurons. Next, morphological analyses of GFP-labelled neurites using the Sholl method revealed a significant reduction in the total neuritic length in neurons overexpressing GluD1^R161H^ or GluD1^T752M^, as compared to control neurons (GFP only) and to neurons overexpressing GluD1^WT^ (control: 598 ± 33 µm, GluD1: 577 ± 25 µm, GluD1^R161H^: 466 ± 21 µm, GluD1^T752M^: 407 ± 21 µm; n=41, 44, 42, 44 neurons, respectively, from 3 cultures in each condition; **Figure 4A**). This was associated with a significant reduction of the number of neuritic branches in neurons transfected with GluD1^R161H^ or GluD1^T752M^, as compared to control or GluD1^WT^-transfected neurons (total crossings; control: 184 ± 9, GluD1: 176 ± 9, GluD1^R161H^: 138 ± 6, GluD1^T752M^: 138 ± 6; n=40, 45, 41, 44 neurons, respectively; **Figure 4A**). These findings indicate that the GluD1 R^161^H and T^752^M mutations perturb neurite outgrowth and architecture in neurons.

**Figure 4:**
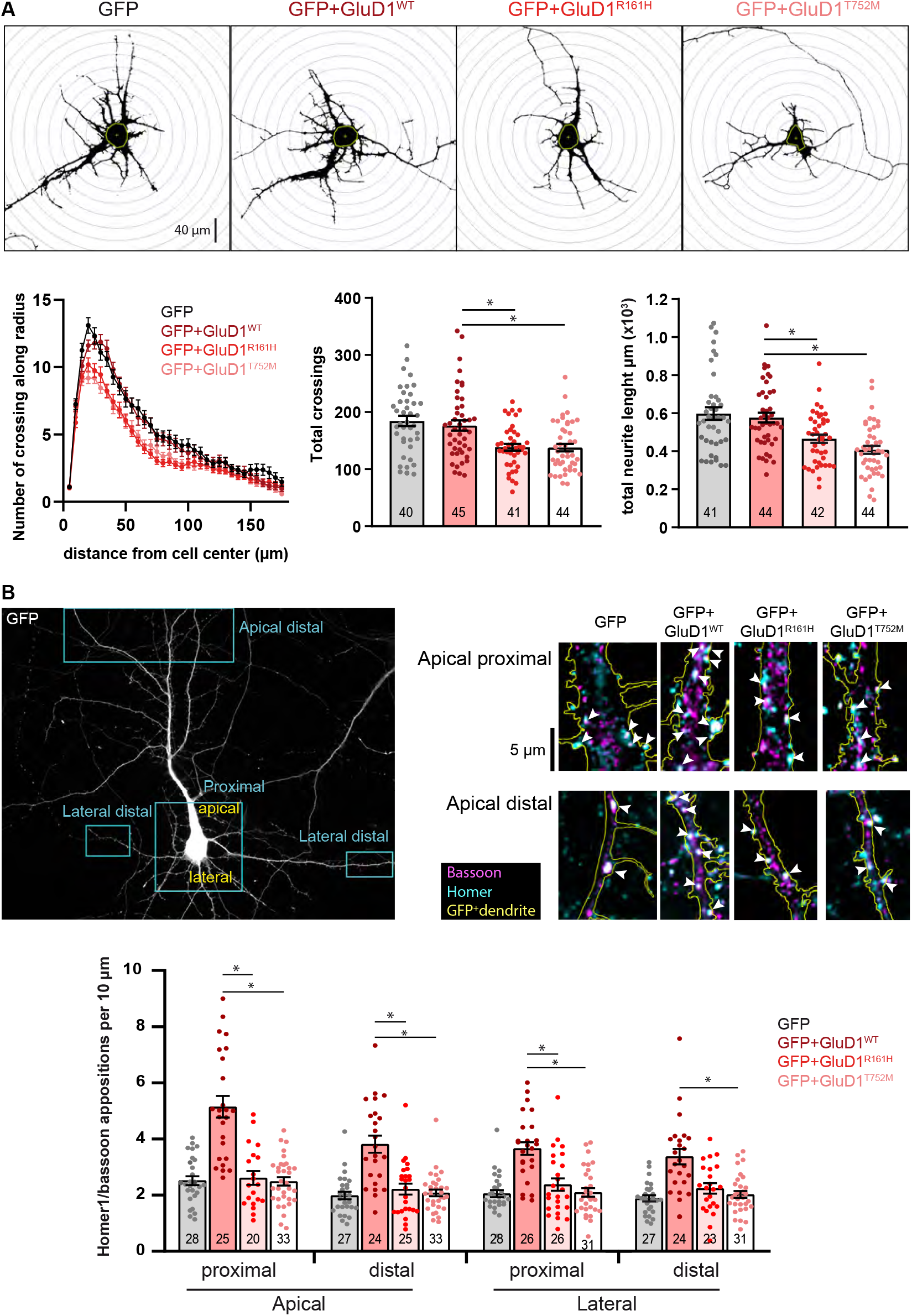
Pathophysiological impact of GluD1^R161H^ and GluD1^T752M^ mutants on neuronal morphology and synaptic density. **(A)** Binary images show GFP fluorescence of cultured hippocampal neurons expressing GFP alone, or GFP and indicated GluD1 variants, after plasmid transfection. Neurites crossing concentric circles centered on each neuron’s soma were counted to quantify neurite ramification. Total neurite length is the sum of all neuritic segments measured for each neuron. Graphs summarize results obtained in n ≥ 40 neurons from 3 cultures in each condition. Note the reduced neuritic length and ramification in neurons expressing GluD1 mutants. **(B)** Squares on the GFP fluorescence picture of a pyramidal-shaped hippocampal neuron in transfected culture (*upper left*) exemplify regions where excitatory putative synapses, revealed by overlap of presynaptic Bassoon and postsynaptic Homer immunostaining on GFP positive dendrites (*upper right*), were counted. The graph shows results obtained in n ≥ 20 pyramidal-shaped hippocampal neurons from 3 cultures in each transfection condition indicated. Note the enhancement of excitatory putative synapse density in GluD1^WT^, but not GluD1 mutants conditions.

GluD1 is present at excitatory synaptic sites (Konno *et al*., 2014; Hepp *et al*., 2015; Benamer *et al*., 2017), and is able to promote the formation of dendritic spines and excitatory synapses (Ryu *et al*., 2012; Gupta *et al*., 2015; Tao *et al*., 2018, Andrews and Dravid, 2021). Since excitatory synapses are localized on dendritic spines of hippocampal principal neurons, we next focused our analyses on spine density and morphology. We found a significant increase in the density of dendritic spines in neurons overexpressing GluD1^WT^ as compared to control neurons (spine number per 10 µm dendritic segment; control: 5.3 ± 0.2, n=23 segments; GluD1^WT^: 6.3 ± 0.2, n=43 segments), consistent with the reported spine-promoting function of GluD1 (Gupta *et al*., 2015). Conversely, neurons overexpressing GluD1^R161H^ exhibited a spine density (5.2 ± 0.2 per 10 µm segment, n=29 segments) similar to that of control neurons (**Figure S8**). We also observed, in the same dendritic sections, that the proportion of immature spines (see Methods and Zagrebelsky *et al*., 2005) was enhanced in GluD1^WT^-transfected neurons, and further increased in GluD1^R161H^-transfected neurons (control: 16.8 ± 1.6 %, GluD1: 22.5 ± 1.0, GluD1^R161H^: 29.6 ± 1.5; **Figure S8**). These results indicate that the R^161^H mutation impairs GluD1 stimulatory effects on dendritic spine formation and maturation. Finally, we evaluated the impact of the GluD1 R^161^H and T^752^M mutations on the density of excitatory synapses by counting overlaps of presynaptic Bassoon and postsynaptic Homer immunolabelling on GFP-expressing dendrites of pyramidal-shaped neurons. As shown in **Figure 4B**, we observed a significantly higher density of putative excitatory synapses on both proximal and distal parts of apical and basal dendrites of GluD1^WT^-transfected neurons compared to control neurons (apical proximal: 5.1 ± 0.4 vs. 2.5 ± 0.2; apical distal: 3.8 ± 0.3 vs. 2.0 ± 0.1; lateral proximal: 3.7 ± 0.2 vs. 2.0 ± 0.1; lateral distal: 3.4 ± 0.3 vs. 1.9 ± 0.1 per 10 µm dendrite of GluD1^WT^ vs. control neurons, respectively; n=25, 24, 26, 24 GluD1^WT^ and n=28, 27, 28, 27 control neurons, respectively, from 3 cultures in each condition), consistent with the synaptogenic function of GluD1 (Ryu *et al*., 2012; Gupta *et al*., 2015, Tao *et al*., 2018). Conversely, the density of excitatory synapses on neurons overexpressing GluD1^R161H^ or GluD1^T752M^ was similar to that on control neurons (apical proximal: 2.6 ± 0.2 and 2.5 ± 0.1; apical distal: 2.2 ± 0.2 and 2.1 ± 0.1; lateral proximal: 2.4 ± 0.2 and 2.1 ± 0.1; lateral distal: 2.2 ± 0.2 and 2.0 ± 0.1 per 10 µm dendrite of GluD1^R161H^ and GluD1^T752M^ neurons, respectively; n=20, 25, 26, 23 GluD1^R161H^ and n=33, 33, 31, 31 GluD1^T752M^ neurons, respectively, from 3 cultures in each condition). This suggests that the role of GluD1 in the formation and stabilization of excitatory synaptic contacts is critically impaired by the R^161^H and T^752^M mutations, despite cerebellin binding, and thus trans-synaptic scaffolding, being preserved in the GluD1 mutants.

The results of morphological analyses collectively indicate that neurite outgrowth, architecture, spine density and maturation, and excitatory synapse density are impaired by the GluD1 R^161^H and T^752^M mutations. Given the widespread distribution of GluD1 (Konno *et al*., 2014; Hepp *et al*., 2015), the R^161^H and T^752^M mutations are thus likely to affect critically the formation and function of brain networks.

## Discussion

We report genetic and functional evidence of the association between homozygous missense variants p.Arg161His and p.Thr752Met in the *GRID1* gene encoding the GluD1 receptor, and disease phenotypes including ID, SPG, and glaucoma, in siblings born to consanguineous parents. Our experimental findings indicate that the p.Arg161His and p.Thr752Met *GRID1* variants impair mGlu1/5 signaling via the ERK pathway, as well as dendritic morphology and excitatory synapse density, in neurons of primary cultures.

### Homozygous *GRID1* variants causing intellectual disability and spastic paraplegia with or without glaucoma

The *GRID1* gene is embedded in the 10q22q23 region, which is subject to recurrent deletions and duplications that cause a broad phenotypic spectrum from healthy status to speech and language delay, and facial dysmorphism (van Bon *et al*., 2011). Several studies have suggested *GRID1* as a candidate gene for neuropsychiatric disorders, based on association of genetic variations in *GRID1* non-coding regions with schizophrenia (Fallin *et al*., 2005; Treutlein *et al*., 2009; Nenadic *et al*., 2012), autism (Griswold *et al*., 2012), risk of bipolar disorder (Zhang *et al*., 2018), and on *GRID1* expression being consistently altered in neuronal precursors and neurons derived from iPS cells of patients with syndromic ID caused by *CDKL5, MECP2* or *FOXG1* gene variants (Livide *et al*., 2015; Patriarchi *et al*., 2016). Moreover, heterozygous missense variants of *GRID1* have been associated with epilepsy (Klassen *et al*., 2011), and with severe undiagnosed developmental disorder (Fitzgerald *et al*., 2015). However, these candidate genetic variations have remained unexplored at the functional level in order to assess their pathogenic contribution. Here, we report the characterization of pathogenic recessive mutations in *GRID1* linked to syndromic ID and SPG without (p.Thr752Met) or with (p.Arg161His) glaucoma, a triple phenotypic association whose genetic bases had not been elucidated previously. Together with earlier descriptions of *GRID2* alterations having extended neurological impact (e.g. Van Schil *et al*., 2015; Ali *et al*., 2017; Grigorenko *et al*., 2022), the present report stresses the importance of the GluD1/2 receptor family in multiple functions of the nervous system, consistent with the widespread expression of both proteins, and with the sensory, behavioral, learning and memory deficits observed in *Grid1*^*−/−*^ mice (Gao *et al*., 2007; Yadav *et al*., 2012; Yadav *et al*., 2013; Konno *et al*., 2014; Hepp *et al*., 2015; Nakamoto *et al*., 2019; Liu *et al*., 2020). Of note, while the two affected sibships reported herein share some phenotypic features and differ with regard to others (**Table 2**), future identification of additional affected individuals will shed more light on the full clinical spectrum of this unique, previously unrecognized disorder and perhaps enable elucidation of possible genotype-phenotype correlations. In this context, inter- and intra-familial phenotypic variability is well-described in numerous inherited neurodevelopmental disorders (e.g. Hanly et al., 2021) and hereditary SPG (Klebe et al., 2015) and might explain some of the observed differences.

The constant association of ID with SPG and glaucoma is rare, as only two affected families have been described (Heijbel and Jagell, 1981, Chenevix-Trench *et al*., 1986), but glaucoma has been mentioned in patients affected with SPG45 (two sisters) and SP75 (one patient), two conditions usually comprising only ID and SPG (Novarino et al., 2014; Lossos et al., 2015). GluD1 is expressed in neurons and at connections with direct relevance for ID: throughout the forebrain, for SPG: in motor cortex and spinal motoneurons, and for glaucoma: in retinal bipolar and ganglion cells, sensory thalamus and superior colliculus (Tolle *et al*., 1993; Brandstatter *et al*., 1997; Jacobs *et al*., 2007; Konno *et al*., 2014; Hepp *et al*., 2015). GluD1 localization at the postsynaptic density, participation in trans-synaptic scaffold, and involvement in glutamatergic transmission and plasticity, point to a role at excitatory synapses (Konno *et al*., 2014; Hepp *et al*., 2015; Benamer *et al*., 2018; Tao *et al*. 2018; Liu *et al*., 2020; Dai *et al*., 2021). Many genetic variants linked to ID concern proteins participating in synaptic function and/or structure (Banerjee *et al*., 2019; Bagni and Zukin, 2019; Lima Caldeira *et al*., 2019). Although most genes identified in SPG do not encode synaptic proteins (Blackstone, 2018; Boutry *et al*., 2019), several SPG-linked variants impact synapses, as illustrated by mutations causing both SPG and ID in the *AP4M1* gene involved in vesicle trafficking of glutamate receptors (Yap *et al*., 2003; Matsuda *et al*., 2008; Abou Jamra *et al*., 2011; Bettencourt *et al*., 2017). Likewise, glaucoma-associated genes identified so far do not encode synaptic proteins (Liu and Allingham, 2017; Trivli *et al*., 2020), but synaptic changes appear to underlie early dysfunction of retinal ganglion cells in this pathology (Agostinone and Di Polo, 2015). Hence, the *GRID1* p.Arg161His and p.Thr752Met variants are rare examples of genetic alteration in a synaptic protein causing ID and SPG with or without glaucoma, but the existence of such mutations is consistent with synaptic impairments occurring in all three pathologies.

### The GluD1 R^161^H and T^752^M mutants impair mGlu1-5 signaling

The recessive nature of p.Arg161His and p.Thre752Met *GRID1* variants indicates that the combination of GluD1^R161H^ or GluD1^T752M^ with GluD1^WT^ subunits leads to functional GluD1 tetramers (Burada *et al*., 2020; Burada *et al*., 2021). Consistent with our molecular modeling study of the GluD1^R161H^ or GluD1^T752M^ mutants predicting only discrete and localized changes in GluD1 structure, we did not observe conspicuous effects of R^161^H and T^752^M mutations on GluD1 expression, stability, membrane targeting, dendritic spine sorting, and association with mGlu1. We also found that both GluD1 mutants bind cerebellin, suggesting that the trans-synaptic scaffolding function is preserved in the mutants. Conversely, our molecular modeling results suggest that R^161^H and T^752^M mutations can affect GluD1 function by altering the transduction of ligand binding to transmembrane/intracellular signaling of this receptor (Yuzaki and Aricescu, 2017; Burada *et al*., 2021; Dai *et al*., 2021; Carrillo *et al*., 2022). We thus searched for alteration of mGlu1/5 signaling, which involves GluD1, is impaired in *Grid1*^*−/−*^ mice, and whose dysregulation at the level of non-canonical pathways tightly relates to ID (D’Antoni *et al*., 2014; Suryavanshi *et al*., 2016; Stoppel *et al*., 2017; Benamer *et al*., 2018; Wilkerson *et al*., 2018). We found that ERK stimulation by the mGlu1/5-GluD1 complex is hampered by the GluD1 R^161^H and T^752^M mutations. ERK proteins are involved in the control of neurite growth and maintenance, and of synapse formation and plasticity (Impey *et al*., 1999; Sweatt, 2001; Davis and Laroche, 2006; Polleux and Snider, 2010; Lavoie *et al*., 2020). Given the high sensitivity of the corticospinal tract to changes in ERK signaling level (Xing *et al*., 2016), and the importance of mGlu1/5 for the excitability of retinal ganglion cells and their connectivity to thalamic targets (Narushima *et al*., 2016; Li *et al*., 2017), dysregulation of mGlu1/5 signaling by GluD1 mutants may contribute to corticospinal axons and optic nerve damage, thus to SPG and glaucoma. This suggests that part of the pathogenic impact of GluD1 R^161^H and T^752^M mutations stems from impaired signaling of the mGlu1/5-GluD1 complex. Nonetheless, recent studies showing the involvement of GluD1 in α1-adrenoceptor signaling and in control of synaptic AMPA/NMDA ratio (Gantz *et al*., 2020; Dai *et al*., 2021), and the cooperative gating of GluD1 channels by cerebellin and D-serine (Carrillo *et al*., 2022), suggest that the GluD1 R^161^H and T^752^M mutations may have additional deleterious effects on the nervous system through dysregulation of other signaling pathways.

### The GluD1 R^161^H and T^752^M mutants impair dendrite morphology and excitatory synapse density

Consistent with the role of GluD1 at excitatory synapses (Ryu *et al*., 2012; Konno *et al*., 2014; Gupta *et al*., 2015; Hepp *et al*., 2015; Benamer *et al*., 2018; Tao *et al*. 2018; Liu *et al*., 2020; Dai *et al*., 2021), we found that dendrite outgrowth, architecture, spine density and maturation, and synapse density are impaired by the GluD1 p.Arg161His and p.Thr752Met mutations. These alterations occurred despite cerebellin binding, thus trans-synaptic scaffolding, being essentially preserved in both GluD1 mutants, confirming that transmembrane signaling by GluD1 is essential to its role in the formation and regulation of excitatory synapses (Tao *et al*. 2018; Dai *et al*., 2021). It is established that loss of expression or function of GluD1 impairs glutamatergic synapses on both spiny and aspiny neurons of diverse excitatory or inhibitory types in the forebrain, midbrain and cerebellum (Konno *et al*., 2014; Gupta *et al*., 2015; Benamer *et al*., 2018; Tao *et al*., 2018; Liu *et al*., 2020; Andrews and Dravid, 2021; Dai *et al*., 2021). This suggests that dendritic and synaptic alterations observed in hippocampal neurons expressing the GluD1 mutants can also affect other GluD1-expressing neuron types relevant for ID, SPG and glaucoma in the forebrain, spinal cord, and retina. Indeed, the tight link between excitatory synapse dysfunction and ID is well documented (Banerjee *et al*., 2019; Bagni and Zukin, 2019; Lima Caldeira *et al*., 2019). Moreover, as discussed above, dysfunction of input and output synapses of corticospinal and retinal ganglion neurons, is likely to have a pathophysiological impact on the corticopinal tract and the optic nerve that may lead to SPG and glaucoma, respectively.

In conclusion, we report the first pathogenic variants of the *GRID1* gene in patients presenting with ID and SPG with or without glaucoma. We provide evidence that the *GRID1* p.Arg161His and Thr752Met mutations impair mGlu1/5 signaling, dendrite outgrowth, architecture, spine density and maturation, and synapse density. Although the present study does not exhaust the possible pathophysiological effects of GluD1^R161H^ and GluD1^T752M^ mutants, our observations demonstrate that their expression has deleterious consequences on neurons and circuits that can cause ID, SPG and glaucoma.

## Data Availability

All data produced in the present study are available upon reasonable request to the authors

## Acknowledgements

The authors would like to thank the patients and their families for their participation and kind cooperation, Pr. Jamel Chelly and Nicolas Lebrun for fruitful discussions and advice during the first steps of the study, Agnès Rastetter for help in genetic analyses, the Bioinformatics Platform of Imagine Institute (linkage analyses), and the IBPS Cell Imaging and Animal Facilities. The study received support and funding from the University of Tours, INSERM, Fondation Maladies Rares (to AT), European Union FP7 Project GENCODYS (to FL, grant number 241995), Association pour le Développement de la Neurogénétique (to FL), the French Agency for Research (to LT, ANR-16-CE16-0014-01) and the Jerôme Lejeune foundation (to RH, grant numbers 1693 and 1959).

## Figure legends

**Supplementary Figure 1:**
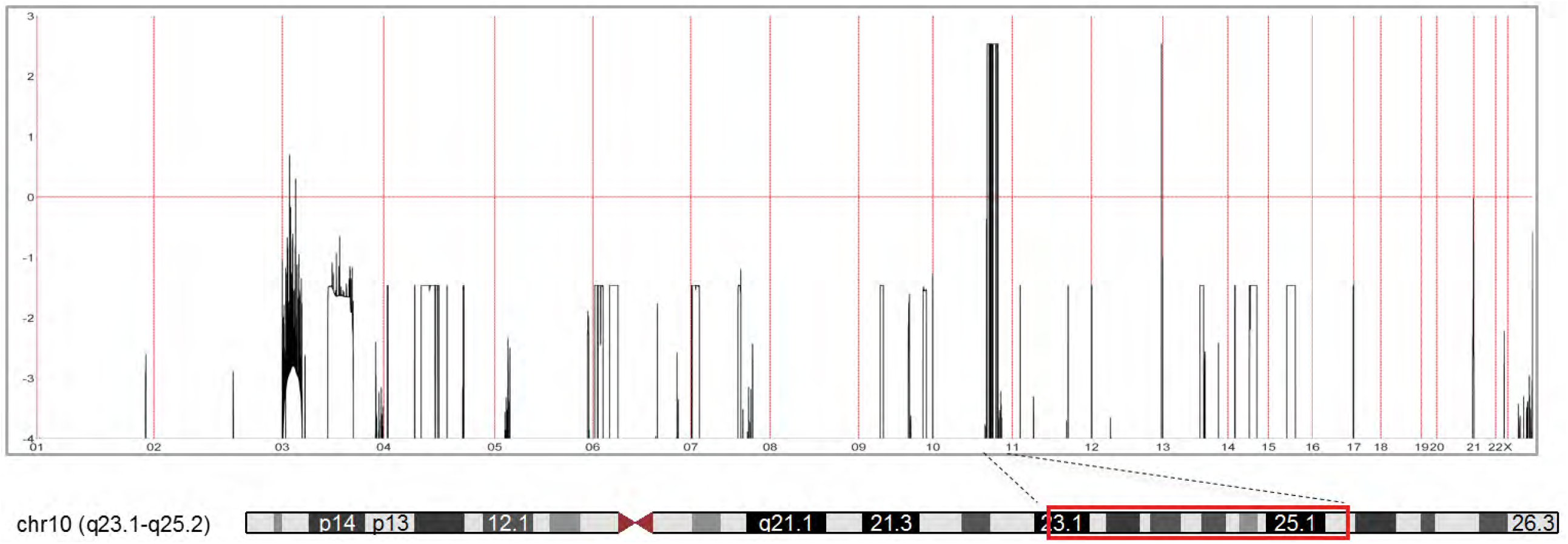
Genome-wide homozygosity mapping data in family. **A** The Y axis represents the LOD score and the X axis represents the genetic distance (chromosomes). Two regions have a maximum LOD score, the largest within chromosome 10 (encompassing *GRID1*) and the second one within the telomeric region of the long arm of chromosome 12.

**Supplementary Figure 2:**
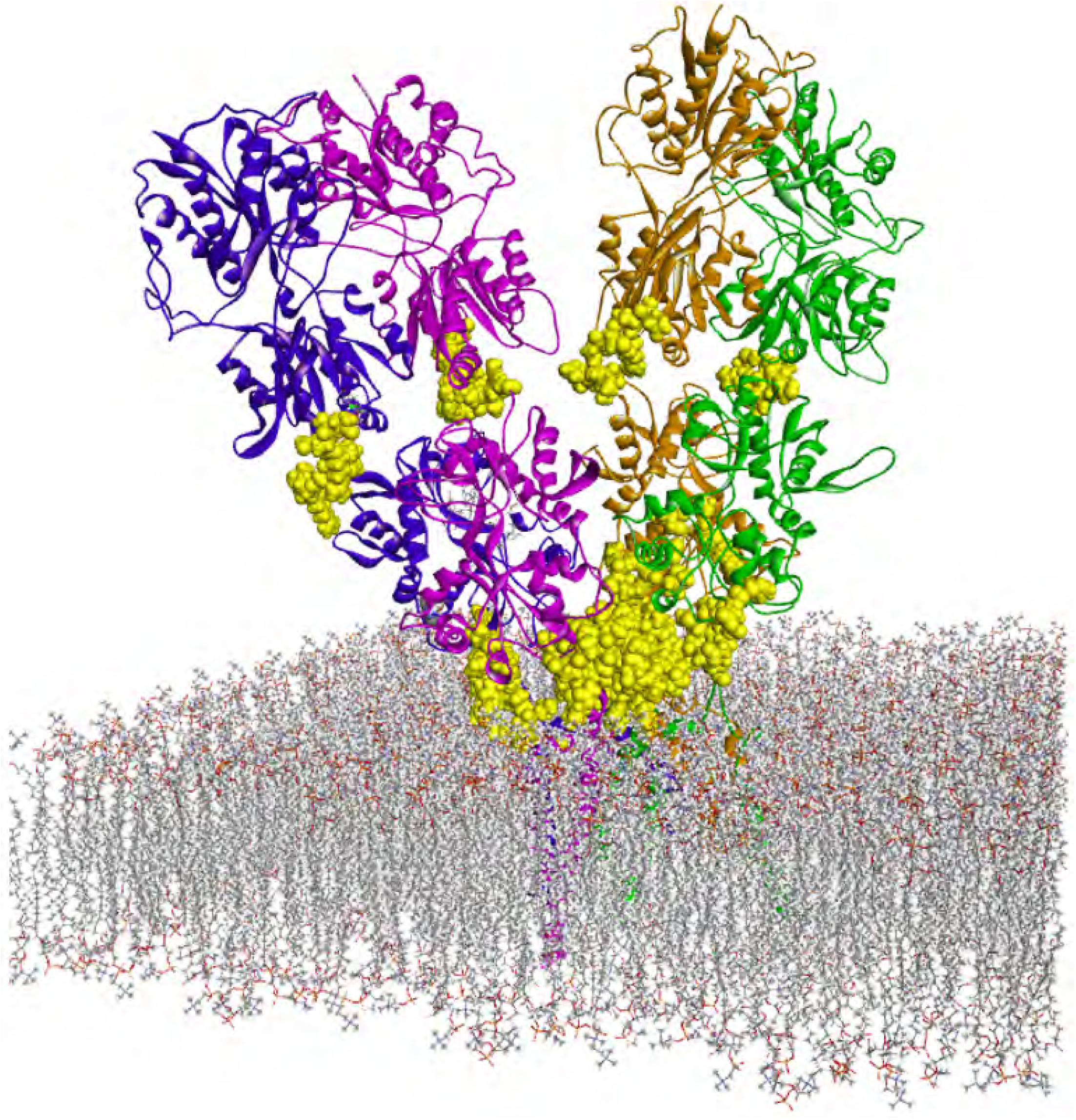
Arrangement of extracellular domains in the GluD1 homotetramer. Modelled complete structure of the GluD1 homotetramer derived from Burada *et al*. (2020) and including the newly generated 3D loops (yellow) between ATD and LBD, and between LBD and transmembrane domains.

**Supplementary Figure 3:**
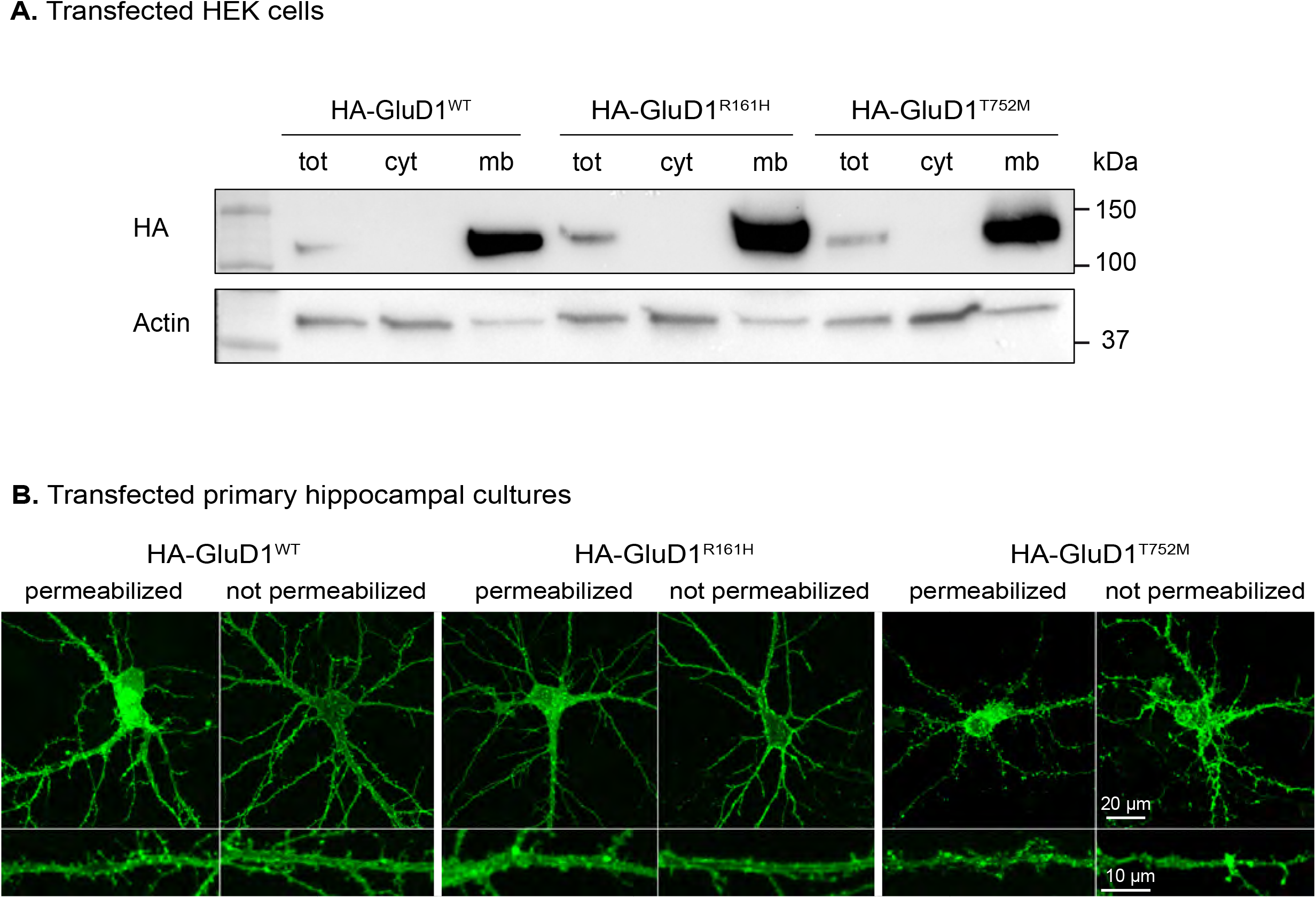
The R^161^H and T^752^M mutations do not hamper the expression and trafficking to the plasma membrane of GluD1. **(A)** Immunoblots of protein lysates (total, cytosolic and membranes fractions) extracted from HEK cells expressing HA-GluD1^WT^, HA-GluD1^R161H^, or HA-GluD1^T752M^ (predicted molecular weight 110 kDa). Beta-actin was used as protein loading control. Similar results were obtained in n=3 independent experiments in each condition. **(B)** Confocal microscopy images of spiny hippocampal neurons from primary cell cultures transfected with plasmids encoding HA-GluD1^WT^, HA-GluD1^R161H^, or HA-GluD1^T752M^, and revealed using anti-HA immunostaining. A zoomed area of a dendritic section is presented for each condition. Similar results were obtained on at least 9 neurons examined from n=3 independent experiments in each condition.

**Supplementary Figure 4:**
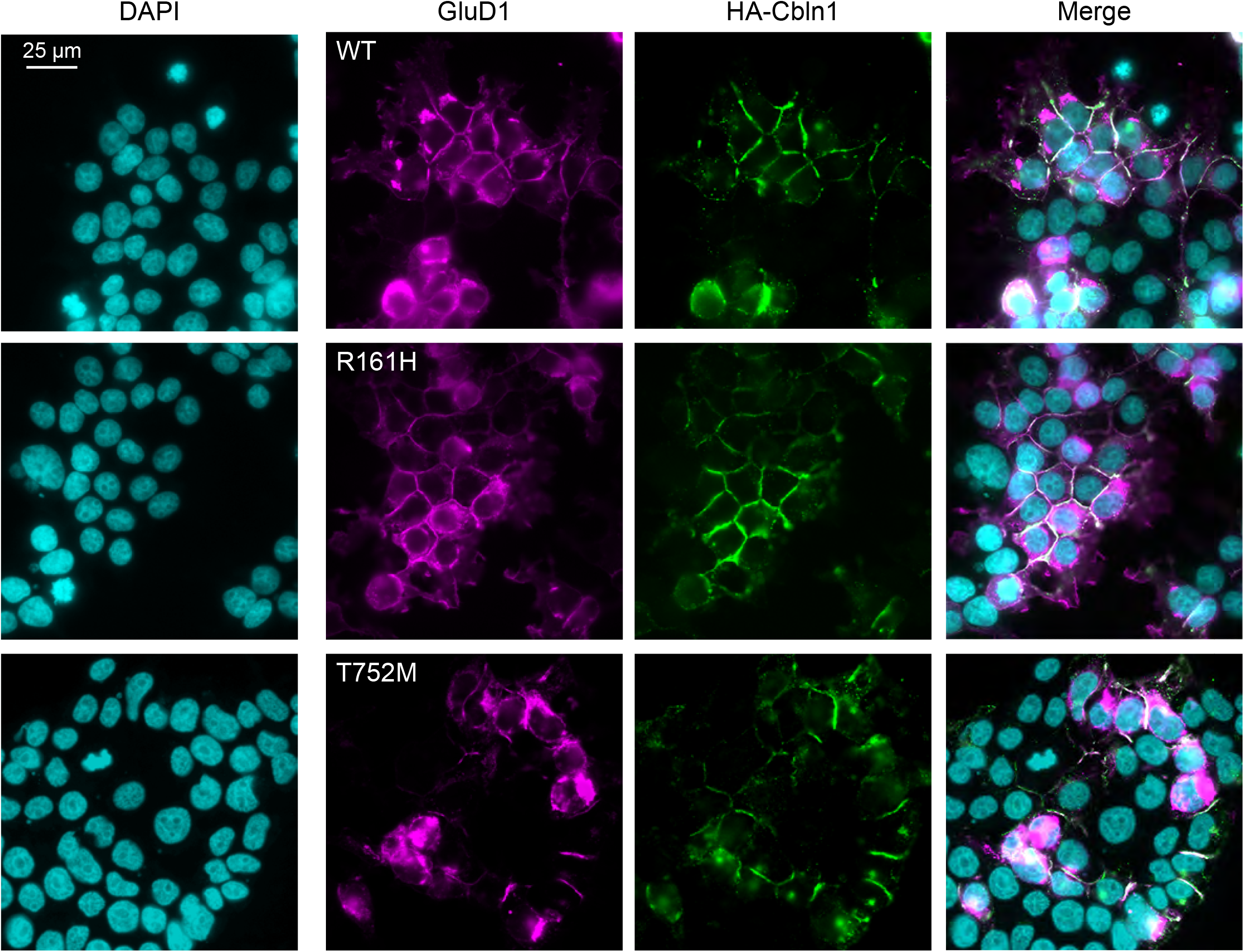
The GluD1 R^161^H and T^752^M mutations do not preclude cerebellin binding. Fluorescence pictures of HEK cells expressing GluD1^WT^, GluD1^R161H^, or GluD1^T752M^ and incubated with HA-tagged cerebellin (HA-Cbln1) prior to fixation, immunolabelling of GluD1 and HA-Cbln1, and nuclear staining with DAPI. Note that HA-Cbln1 immunostaining was similar for GluD1^WT^-, GluD1^R161H^-, and GluD1^T752M^-expressing cells, and that HA-Cbln1 binding was not detected on GluD1^WT^-, GluD1^R161H^-, and GluD1^T752M^-negative cells. Similar results were obtained in n=3 independent experiments in each condition.

**Supplementary Figure 5:**
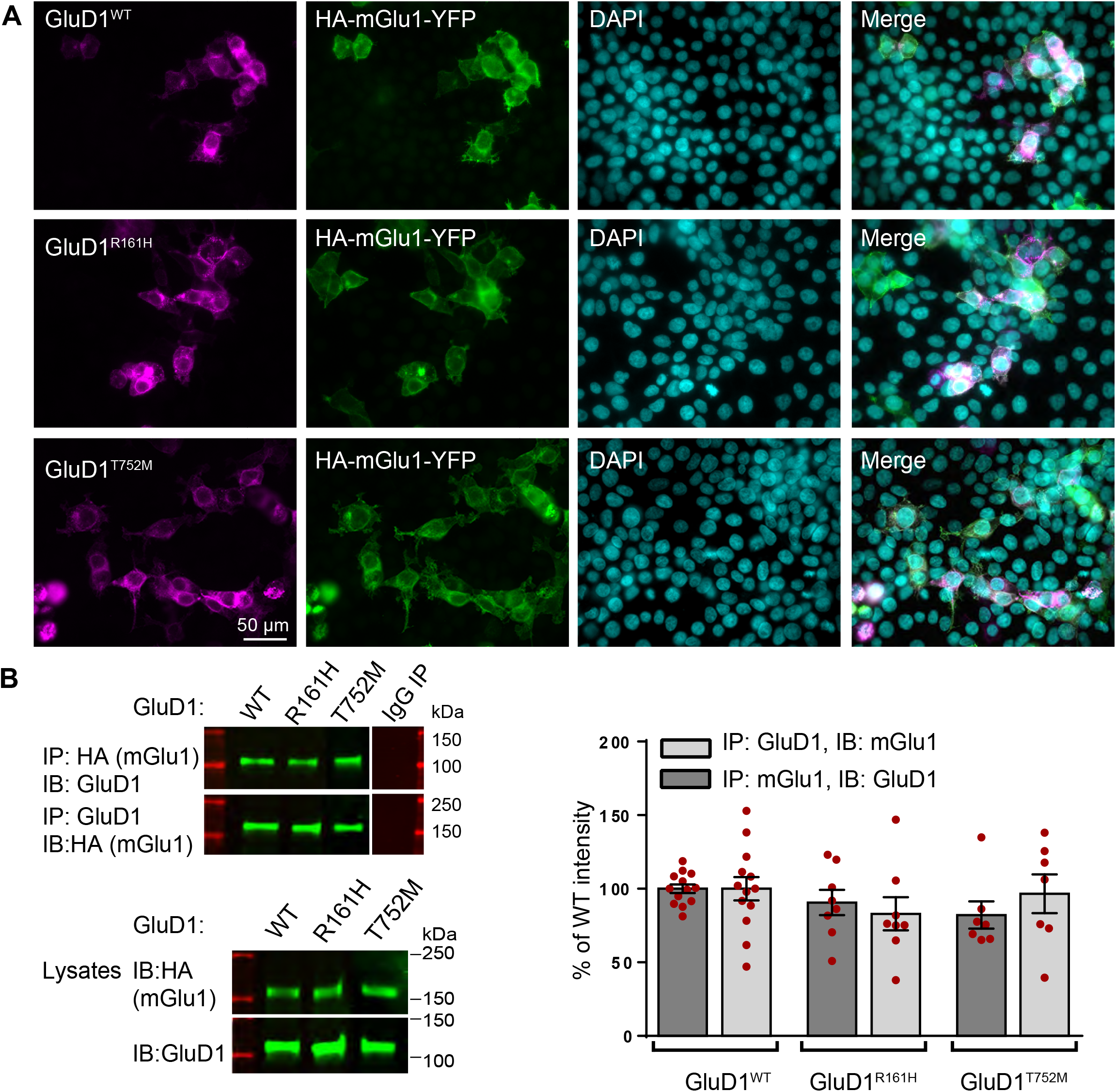
The GluD1 R^161^H and T^752^M mutations do not hamper mGlu1-GluD1 physical interaction. **(A)** Fluorescence pictures of HEK cells co-expressing GluD1^WT^, GluD1^R161H^, or GluD1^T752M^ and HA-tagged mGlu1. **(B)** *Left panel*: HEK cell lysates were subjected to immunoprecipitation (IP) with indicated antibodies against GluD1, HA-tagged mGlu1, or with a control antibody (IgG). Immunoblotting (IB) of IP eluates or cell lysates were probed using indicated antibodies. *Right panel*: The bar graph summarizes results of 3 experiments performed in duplicate for each mGlu1+GluD1^WT^/GluD1^R161H^/GluD1^T752M^ combination. The mean intensity of the bands GluD1^WT^ pulled down by mGlu1 and of the bands mGlu1 pulled down by GluD1^WT^ was normalized to 100%. Results of experiments involving GluD1 mutants are expressed as % of results involving GluD1^WT^.

**Supplementary Figure 6:**
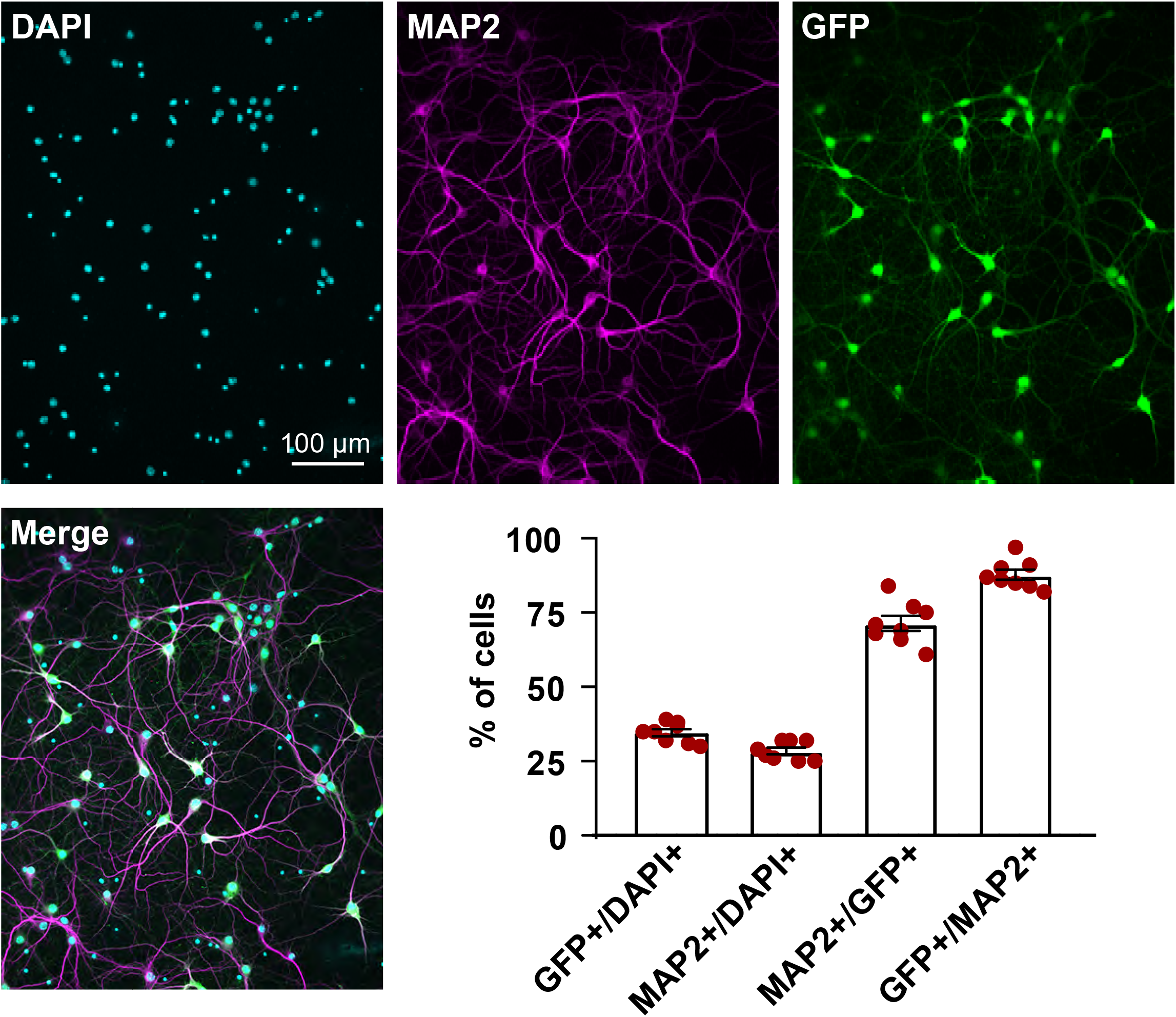
Lentiviral transfer of GluD1-ires-GFP in neurons of primary cortical cell cultures from *Grid1*−/− mice. Fluorescence pictures of a primary cortical cell culture transduced with a lentivirus co-expressing GluD1 and GFP, and processed for immunolabelling of GFP and the neuronal marker MAP2, and for nuclear staining with DAPI. The graph summarizes results obtained on 6823 DAPI-positive cells from 2 cultures, 8 coverslips, 5 area analyzed per coverslip. Among DAPI-positive cells, 35 ± 1 % were GFP-positive, and 28 ± 1 % were MAP2-positive. Note that 71 ± 3 % of GFP-positive cells were Map2-positive, and that 88 ± 2 % of MAP2-positive cells were GFP-positive, showing that GluD1-expressing lentiviruses preferentially and efficiently transduced neurons.

**Supplementary Figure 7:**
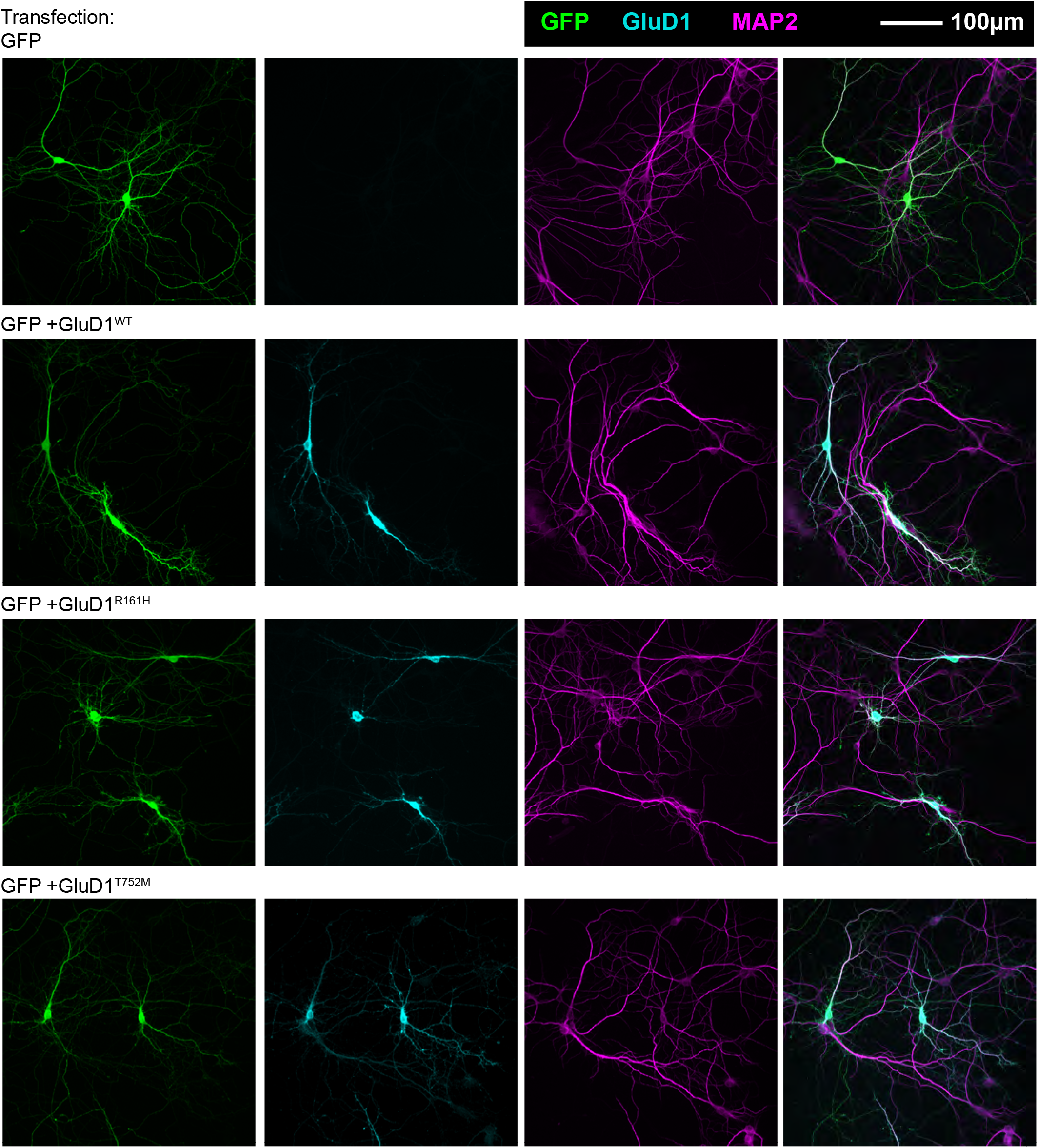
Expression of transfected GluD1^WT^ and GluD1 mutants is largely superior to endogenous GluD1 in *Grid1*^+/+^ hippocampal neurons. Fluorescence pictures showing hippocampal neurons in culture immunostained for GFP, GluD1 and the neuronal marker MAP2 after transfection of indicated plasmids. Note that transfected cells express MAP2, and that expression of transfected GluD1 is far superior to endogenous GluD1, as evidenced by the very faint immunostaining of non-transfected neurons.

**Supplementary Figure 8:**
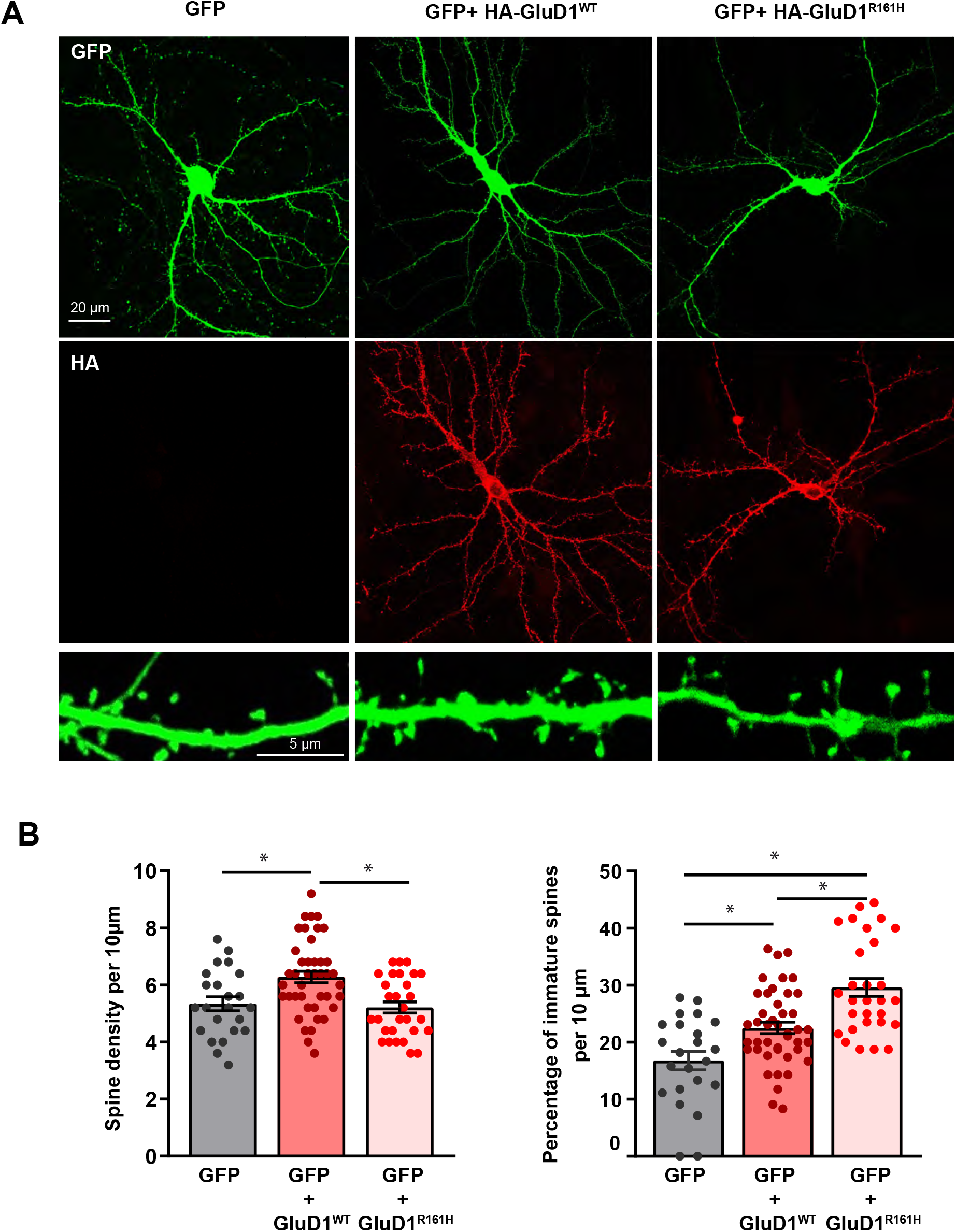
Pathophysiological impact of the GluD1^R161H^ mutant on dendritic spines. **(A)** *Upper panels*: Confocal microscopy images of cultured hippocampal neurons expressing GFP alone, or both GFP and HA-GluD1^WT^ or HA-GluD1^R161H^, after plasmid transfection. All GFP-expressing neurons examined after co-transfection also expressed either HA-GluD1^WT^ (n=64) or HA-GluD1^R161H^, (n=64) as shown by anti-HA immunostaining of 5 independent culture transfections. *Lower panels*: Confocal microscopy images of spiny dendritic sections of hippocampal neurons transfected as indicated. **(B)** Graphs summarizing results of spine density and morphology analyses performed on n=23 (GFP), 43 (GFP+ HA-GluD1^WT^), and 29 (GFP+ HA-GluD1^R161H^) segments of spiny dendrites from at least 3 independent cultures in each condition. Immature spines comprise thin-long and filopodia-shaped spines, as opposed to mushroom-shaped and stubby mature spines.

## Notes

### Competing Interest Statement

The authors have declared no competing interest.

### Funding Statement

The study was funding by the University of Tours, INSERM, Fondation Maladies Rares (to AT), European Union FP7 Project GENCODYS (to FL, grant number 241995), Association pour le Developpement de la Neurogenetique (to FL), the French Agency for Research (to LT, ANR-16-CE16-0014-01) and the Jerome Lejeune Foundation (to RH, grant numbers 1693 and 1959).

### Author Declarations

Ethics committee/IRB)-of the Centre Hospitalier Universitaire de Tours (France) and of the Hadassah Medical Center (Israel) gave ethical approval for this work

## References

Abou Jamra R, Philippe O, Raas-Rothschild A, Eck SH, Graf E, Buchert R, Borck G, Ekici A, Brockschmidt FF, Nöthen MM, Munnich A, Strom TM, Reis A, Colleaux L. Adaptor protein complex 4 deficiency causes severe autosomal-recessive intellectual disability, progressive spastic paraplegia, shy character, and short stature. Am J Hum Genet. 2011 Jun 10;88(6):788–795.

Ady V, Perroy J, Tricoire L, Piochon C, Dadak S, Chen X, Dusart I, Fagni L, Lambolez B, Levenes C. Type 1 metabotropic glutamate receptors (mGlu1) trigger the gating of GluD2 delta glutamate receptors. EMBO Rep. 2014 Jan;15(1):103–9.

Agostinone J, Di Polo A. Retinal ganglion cell dendrite pathology and synapse loss: Implications for glaucoma. Prog Brain Res. 2015;220:199–216.

Ali Z, Zulfiqar S, Klar J, Wikström J, Ullah F, Khan A, Abdullah U, Baig S, Dahl N. Homozygous GRID2 missense mutation predicts a shift in the D-serine binding domain of GluD2 in a case with generalized brain atrophy and unusual clinical features. BMC Med Genet. 2017 Dec 6;18(1):144.

Andrews PC, Dravid SM. An emerging map of glutamate delta 1 receptors in the forebrain. Neuropharmacology. 2021 Jul 1;192:108587.

Araki K, Meguro H, Kushiya E, Takayama C, Inoue Y, Mishina M. Selective expression of the glutamate receptor channel delta 2 subunit in cerebellar Purkinje cells. Biochem Biophys Res Commun 1993; 197: 1267–76.

Bagni C, Zukin RS. A Synaptic Perspective of Fragile X Syndrome and Autism Spectrum Disorders. Neuron. 2019 Mar 20;101(6):1070–1088.

Banerjee A, Miller MT, Li K, Sur M, Kaufmann WE. Towards a better diagnosis and treatment of Rett syndrome: a model synaptic disorder. Brain. 2019 Feb1;142(2):239–248.

Benamer N, Marti F, Lujan R, Hepp R, Aubier TG, Dupin AAM, Frébourg G, Pons S, Maskos U, Faure P, Hay YA, Lambolez B, Tricoire L. GluD1, linked to schizophrenia, controls the burst firing of dopamine neurons. Mol Psychiatry. 2018 Mar;23(3):691–700.

Bettencourt C, Salpietro V, Efthymiou S, Chelban V, Hughes D, Pittman AM, Federoff M, Bourinaris T, Spilioti M, Deretzi G, Kalantzakou T, Houlden H, Singleton AB, Xiromerisiou G. Genotype-phenotype correlations and expansion of the molecular spectrum of AP4M1-related hereditary spastic paraplegia. Orphanet J Rare Dis. 2017 Nov 2;12(1):172.

Blackstone C. Converging cellular themes for the hereditary spastic paraplegias. Curr Opin Neurobiol. 2018 Aug;51:139–146.

Brandstätter JH, Koulen P, Wässle H. Selective synaptic distribution of kainate receptor subunits in the two plexiform layers of the rat retina. J Neurosci. 1997 Dec 1;17(23):9298–307.

Burada AP, Vinnakota R, Kumar J. Cryo-EM structures of the ionotropic glutamate receptor GluD1 reveal a non-swapped architecture. Nat Struct Mol Biol. 2020 Jan;27(1):84–91.

Burada AP, Vinnakota R, Lambolez B, Tricoire L, Kumar J. Structural biology of ionotropic glutamate delta receptors and their crosstalk with metabotropic glutamate receptors. Neuropharmacology. 2021 Jun 25:108683.

Carrillo E, Gonzalez CU, Berka V, Jayaraman V. Delta glutamate receptors are functional glycine-and ?-serine-gated cation channels in situ. Sci Adv. 2021 Dec 24;7(52):eabk2200.

Chenevix-Trench G, Leshner R, Mamunes P. Spastic paresis, glaucoma and mental retardation--a probable autosomal recessive syndrome? Clin Genet 1986; 30: 416–21.

Cooper GM, Coe BP, Girirajan S, Rosenfeld JA, Vu TH, Baker C, Williams C, Stalker H, Hamid R, Hannig V, Abdel-Hamid H, Bader P, McCracken E, Niyazov D, Leppig K, Thiese H, Hummel M, Alexander N, Gorski J, Kussmann J, Shashi V, Johnson K, Rehder C, Ballif BC, Shaffer LG, Eichler EE. A copy number variation morbidity map of developmental delay. Nat Genet. 2011 Aug 14;43(9):838–46.

Coutelier M, Burglen L, Mundwiller E, Abada-Bendib M, Rodriguez D, Chantot-Bastaraud S, Rougeot C, Cournelle MA, Milh M, Toutain A, Bacq D, Meyer V, Afenjar A, Deleuze JF, Brice A, Héron D, Stevanin G, Durr A. GRID2 mutations span from congenital to mild adult-onset cerebellar ataxia. Neurology. 2015 Apr 28;84(17):1751–9.

D’Antoni S, Spatuzza M, Bonaccorso CM, Musumeci SA, Ciranna L, Nicoletti F, Huber KM, Catania MV. Dysregulation of group-I metabotropic glutamate (mGlu) receptor mediated signalling in disorders associated with Intellectual Disability and Autism. Neurosci Biobehav Rev. 2014 Oct;46 Pt 2(Pt 2):228–41.

Dai J, Patzke C, Liakath-Ali K, Seigneur E, Südhof TC. GluD1 is a signal transduction device disguised as an ionotropic receptor. Nature. 2021 Jun 16.

Davis S, Laroche S. Mitogen-activated protein kinase/extracellular regulated kinase signalling and memory stabilization: a review. Genes Brain Behav. 2006;5 Suppl 2:61–72.

Dhers L, Pietrancosta N, Ducassou L, Ramassamy B, Dairou J, Jaouen M, André F, Mansuy D, Boucher JL. Spectral and 3D model studies of the interaction of orphan human cytochrome P450 2U1 with substrates and ligands. Biochim Biophys Acta Gen Subj. 2017 Jan;1861(1 Pt A):3144–3153.

Drobac E, Tricoire L, Chaffotte AF, Guiot E, Lambolez B. Calcium imaging in single neurons from brain slices using bioluminescent reporters. J Neurosci Res. 2010 Mar;88(4):695–711.

Ducassou L, Jonasson G, Dhers L, Pietrancosta N, Ramassamy B, Xu-Li Y, Loriot MA, Beaune P, Bertho G, Lombard M, Mansuy D, André F, Boucher JL. Expression in yeast, new substrates, and construction of a first 3D model of human orphan cytochrome P450 2U1: Interpretation of substrate hydroxylation regioselectivity from docking studies. Biochim Biophys Acta. 2015 Jul;1850(7):1426–37.

Elegheert J, Kakegawa W, Clay JE, Shanks NF, Behiels E, Matsuda K, Kohda K, Miura E, Rossmann M, Mitakidis N, Motohashi J, Chang VT, Siebold C, Greger IH, Nakagawa T, Yuzaki M, Aricescu AR. Structural basis for integration of GluD receptors within synaptic organizer complexes. Science. 2016 Jul 15;353(6296):295–9.

Ellison JW, Rosenfeld JA, Shaffer LG. Genetic basis of intellectual disability. Annu Rev Med 2013; 64: 441–50.

Elsayed LEO, Eltazi IZ, Ahmed AE, Stevanin G. Insights into Clinical, Genetic, and Pathological Aspects of Hereditary Spastic Paraplegias: A Comprehensive Overview. Front Mol Biosci. 2021 Nov 26;8:690899.

Fallin MD, Lasseter VK, Avramopoulos D, Nicodemus KK, Wolyniec PS, McGrath JA, Steel G, Nestadt G, Liang KY, Huganir RL, Valle D, Pulver AE. Bipolar I disorder and schizophrenia: a 440-single-nucleotide polymorphism screen of 64 candidate genes among Ashkenazi Jewish case-parent trios. Am J Hum Genet. 2005 Dec;77(6):918–36.

Ferreira TA, Blackman AV, Oyrer J, Jayabal S, Chung AJ, Watt AJ, Sjöström PJ, van Meyel DJ. Neuronal morphometry directly from bitmap images. Nat Methods. 2014 Oct;11(10):982–4.

Fitzgerald TW, Gerety SS, Jones WD, van Kogelenberg M, King DA, McRae J, Morley KI et al. Deciphering Developmental Disorders Study. Large-scale discovery of novel genetic causes of developmental disorders. Nature 2015; 519: 223–8.

Gantz SC, Moussawi K, Hake HS. Delta glutamate receptor conductance drives excitation of mouse dorsal raphe neurons. Elife. 2020 Apr 1;9:e56054.

Gao J, Maison SF, Wu X, Hirose K, Jones SM, Bayazitov I, Tian Y, Mittleman G, Matthews DB, Zakharenko SS, Liberman MC, Zuo J. Orphan glutamate receptor delta1 subunit required for high-frequency hearing. Mol Cell Biol. 2007 Jun;27(12):4500–12.

Glessner JT, Wang K, Cai G, Korvatska O, Kim CE, Wood S, et al. Autism genome-wide copy number variation reveals ubiquitin and neuronal genes. Nature 2009; 459: 569–73.

Grigorenko AP, Protasova MS, Lisenkova AA, Reshetov DA, Andreeva TV, Garcias GL, Martino Roth MDG, Papassotiropoulos A, Rogaev EI. Neurodevelopmental Syndrome with Intellectual Disability, Speech Impairment, and Quadrupedia Is Associated with Glutamate Receptor Delta 2 Gene Defect. Cells. 2022 Jan 25;11(3):400

Griswold AJ, Ma D, Cukier HN, Nations LD, Schmidt MA, Chung RH, et al. Evaluation of copy number variations reveals novel candidate genes in autism spectrum disorder-associated pathways. Hum Mol Genet 2012; 21: 3513–23.

Guo S-Z, Huang K, Shi Y-Y, Tang W, Zhou J, Feng G-Y, et al. A case-control association study between the GRID1 gene and schizophrenia in the Chinese Northern Han population. Schizophr Res 2007; 93: 385–90.

Gupta SC, Yadav R, Pavuluri R, Morley BJ, Stairs DJ, Dravid SM. Essential role of GluD1 in dendritic spine development and GluN2B to GluN2A NMDAR subunit switch in the cortex and hippocampus reveals ability of GluN2B inhibition in correcting hyperconnectivity. Neuropharmacology 2015; 93: 274–84.

Hanly C, Shah H, Au PYB, Murias K. Description of neurodevelopmental phenotypes associated with 10 genetic neurodevelopmental disorders: A scoping review. Clin Genet 2021; 99: 335–46.

Heijbel J, Jagell S. Spastic paraplegia, glaucoma and mental retardation--in three siblings. A new genetic syndrome. Hereditas 1981; 94: 203–7.

Hepp R, Hay YA, Aguado C, Lujan R, Dauphinot L, Potier MC, et al. Glutamate receptors of the delta family are widely expressed in the adult brain. Brain Struct Funct 2015; 220: 2797–815.

Hills LB, Masri A, Konno K, Kakegawa W, Lam A-TN, Lim-Melia E, et al. Deletions in GRID2 lead to a recessive syndrome of cerebellar ataxia and tonic upgaze in humans. Neurology 2013; 81: 1378–86.

Impey S, Obrietan K, Storm DR. Making new connections: role of ERK/MAP kinase signaling in neuronal plasticity. Neuron. 1999 May;23(1):11–4.

Jakobs TC, Ben Y, Masland RH. Expression of mRNA for glutamate receptor subunits distinguishes the major classes of retinal neurons, but is less specific for individual cell types. Mol Vis. 2007 Jun 18;13:933–48.

Jo, S.; Kim, T.; Iyer, V. G.; Im, W. CHARMM-GUI: A Web-Based Graphical User Interface for CHARMM. J. Comput. Chem. 2008, 29, 1859– 1865, DOI: 10.1002/jcc.20945

Klassen T, Davis C, Goldman A, Burgess D, Chen T, Wheeler D, et al. Exome sequencing of ion channel genes reveals complex profiles confounding personal risk assessment in epilepsy. Cell 2011; 145: 1036–48.

Klebe S, Stevanin G, Depienne C. Clinical and genetic heterogeneity in hereditary spastic paraplegias: from SPG1 to SPG72 and still counting. Rev Neurol (Paris) 2015; 171: 505–30.

Konno K, Matsuda K, Nakamoto C, Uchigashima M, Miyazaki T, Yamasaki M, et al. Enriched expression of GluD1 in higher brain regions and its involvement in parallel fiber-interneuron synapse formation in the cerebellum. J Neurosci 2014; 34: 7412–24.

Kristensen AS, Hansen KB, Naur P, Olsen L, Kurtkaya NL, Dravid SM, Kvist T, Yi F, Pøhlsgaard J, Clausen RP, Gajhede M, Kastrup JS, Traynelis SF. Pharmacology and Structural Analysis of Ligand Binding to the Orthosteric Site of Glutamate-Like GluD2 Receptors. Mol Pharmacol. 2016 Feb;89(2):253–62.

Lavoie H, Gagnon J, Therrien M. ERK signalling: a master regulator of cell behaviour, life and fate. Nat Rev Mol Cell Biol. 2020 Oct;21(10):607–632.

Li Q, Cui P, Miao Y, Gao F, Li XY, Qian WJ, Jiang SX, Wu N, Sun XH, Wang Z. Activation of group I metabotropic glutamate receptors regulates the excitability of rat retinal ganglion cells by suppressing Kir and Ih. Brain Struct Funct. 2017 Mar;222(2):813–830.

Lima Caldeira G, Peça J, Carvalho AL. New insights on synaptic dysfunction in neuropsychiatric disorders. Curr Opin Neurobiol. 2019 Aug;57:62–70.

Liu Y, Allingham RR. Major review: Molecular genetics of primary open-angle glaucoma. Exp Eye Res. 2017 Jul;160:62–84.

Liu J, Shelkar GP, Gandhi PJ, Gawande DY, Hoover A, Villalba RM, Pavuluri R, Smith Y, Dravid SM. Striatal glutamate delta-1 receptor regulates behavioral flexibility and thalamostriatal connectivity. Neurobiol Dis. 2020 Apr;137:104746.

Livide G, Patriarchi T, Amenduni M, Amabile S, Yasui D, Calcagno E, et al. GluD1 is a common altered player in neuronal differentiation from both MECP2-mutated and CDKL5-mutated iPS cells. Eur J Hum Genet 2015; 23: 195–201.

Lossos A, Elazar N, Lerer I, Schueler-Furman O, Fellig Y, Glick B, Zimmerman B.-E, Azulay H, Dotan S, Goldberg S, Gomori J. M, Ponger P, et al. Myelin-associated glycoprotein gene mutation causes Pelizaeus-Merzbacher disease-like disorder. Brain 2015;138: 2521–2536.

Maier A, Klopocki E, Horn D, Tzschach A, Holm T, Meyer R, et al. De novo partial deletion in GRID2 presenting with complicated spastic paraplegia. Muscle Nerve 2014; 49: 289–92.

Matsuda S, Miura E, Matsuda K, Kakegawa W, Kohda K, Watanabe M, Yuzaki M. Accumulation of AMPA receptors in autophagosomes in neuronal axons lacking adaptor protein AP-4. Neuron. 2008 Mar 13;57(5):730–45.

Nakamoto C, Konno K, Miyazaki T, Nakatsukasa E, Natsume R, Abe M, Kawamura M, Fukazawa Y, Shigemoto R, Yamasaki M, Sakimura K, Watanabe M. Expression mapping, quantification, and complex formation of GluD1 and GluD2 glutamate receptors in adult mouse brain. J Comp Neurol. 2020 Apr;528(6):1003–1027.

Narushima M, Uchigashima M, Yagasaki Y, Harada T, Nagumo Y, Uesaka N, Hashimoto K, Aiba A, Watanabe M, Miyata M, Kano M. The Metabotropic Glutamate Receptor Subtype 1 Mediates Experience-Dependent Maintenance of Mature Synaptic Connectivity in the Visual Thalamus. Neuron. 2016 Sep 7;91(5):1097–1109.

Nenadic I, Maitra R, Scherpiet S, Gaser C, Schultz CC, Schachtzabel C, et al. Glutamate receptor δ 1 (GRID1) genetic variation and brain structure in schizophrenia. J Psychiatr Res 2012; 46: 1531–9.

Novarino G, Fenstermaker AG, Zaki MS, Hofree M, Silhavy JL, Heiberg AD, Abdellateef M, Rosti B, Scott E, Mansour L, Masri A, Kayserili H, et al. Exome sequencing links corticospinal motor neuron disease to common neurodegenerative disorders. Science 2014;343: 506–511.

Patriarchi T, Amabile S, Frullanti E, Landucci E, Lo Rizzo C, Ariani F, Costa M, Olimpico F W Hell J M Vaccarino F, Renieri A, Meloni I. Imbalance of excitatory/inhibitory synaptic protein expression in iPSC-derived neurons from FOXG1(+/−) patients and in foxg1(+/−) mice. Eur J Hum Genet. 2016 Jun;24(6):871–80.

Perroy J, Raynaud F, Homburger V, Rousset MC, Telley L, Bockaert J et al. Direct interaction enables cross-talk between ionotropic and group I metabotropic glutamate receptors. J Biol Chem 2008; 283(11): 6799–6805.

Polleux F, Snider W. Initiating and growing an axon. Cold Spring Harb Perspect Biol. 2010 Apr;2(4):a001925.

Ryu K, Yokoyama M, Yamashita M, Hirano T. Induction of excitatory and inhibitory presynaptic differentiation by GluD1. Biochem Biophys Res Commun 2012; 417: 157–61.

Schindelin J, Arganda-Carreras I, Frise E, Kaynig V, Longair M, Pietzsch T, Preibisch S, Rueden C, Saalfeld S, Schmid B, Tinevez JY, White DJ, Hartenstein V, Eliceiri K, Tomancak P, Cardona A. Fiji: an open-source platform for biological-image analysis. Nat Methods. 2012 Jun 28;9(7):676–82.

Schmid SM, Hollmann M. To gate or not to gate: are the delta subunits in the glutamate receptor family functional ion channels? Mol Neurobiol. 2008 Apr-Jun;37(2-3):126–41.

Schneider CA, Rasband WS, Eliceiri KW. NIH Image to ImageJ: 25 years of image analysis. Nat Methods. 2012 Jul;9(7):671–5.

Stoppel LJ, Auerbach BD, Senter RK, Preza AR, Lefkowitz RJ, Bear MF. beta-Arrestin2 Couples Metabotropic Glutamate Receptor 5 to Neuronal Protein Synthesis and Is a Potential Target to Treat Fragile X. Cell Rep, 2017. 18(12): p. 2807–2814.

Suryavanshi PS, Gupta SC, Yadav R, Kesherwani V, Liu J, Dravid SM. Glutamate Delta-1 Receptor Regulates Metabotropic Glutamate Receptor 5 Signaling in the Hippocampus. Mol Pharmacol 2016; 90: 96–105.

Sweatt JD. The neuronal MAP kinase cascade: a biochemical signal integration system subserving synaptic plasticity and memory. J Neurochem. 2001 Jan;76(1):1–10.

Tao W, Díaz-Alonso J, Sheng N, Nicoll RA. Postsynaptic δ1 glutamate receptor assembles and maintains hippocampal synapses via Cbln2 and neurexin. Proc Natl Acad Sci U S A. 2018 Jun 5;115(23):E5373–E5381.

Tölle TR, Berthele A, Zieglgänsberger W, Seeburg PH, Wisden W. The differential expression of 16 NMDA and non-NMDA receptor subunits in the rat spinal cord and in periaqueductal gray. J Neurosci. 1993 Dec;13(12):5009–28.

Traynelis SF, Wollmuth LP, McBain CJ, Menniti FS, Vance KM, Ogden KK, Hansen KB, Yuan H, Myers SJ, Dingledine R. Glutamate receptor ion channels: structure, regulation, and function. Pharmacol Rev. 2010 Sep;62(3):405–96.

Treutlein J, Mühleisen TW, Frank J, Mattheisen M, Herms S, Ludwig KU, et al. Dissection of phenotype reveals possible association between schizophrenia and Glutamate Receptor Delta 1 (GRID1) gene promoter. Schizophr Res 2009; 111: 123–30.

Trivli A, Zervou MI, Goulielmos GN, Spandidos DA, Detorakis ET. Primary open angle glaucoma genetics: The common variants and their clinical associations (Review). Mol Med Rep. 2020 Aug;22(2):1103–1110.

Umesono K, Murakami KK, Thompson CC, Evans RM. Direct repeats as selective response elements for the thyroid hormone, retinoic acid, and vitamin D3 receptors. Cell. 1991 Jun 28;65(7):1255–66.

Ung DC, Iacono G, Méziane H, Blanchard E, Papon MA, Selten M, van Rhijn JR, Montjean R, Rucci J, Martin S, Fleet A, Birling MC, Marouillat S, Roepman R, Selloum M, Lux A, Thépault RA, Hamel P, Mittal K, Vincent JB, Dorseuil O, Stunnenberg HG, Billuart P, Nadif Kasri N, Hérault Y, Laumonnier F. Ptchd1 deficiency induces excitatory synaptic and cognitive dysfunctions in mouse. Mol Psychiatry. 2018 May;23(5):1356–1367.

Utine GE, Haliloglu G, Salanci B, Çetinkaya A, Kiper PÖ, Alanay Y, et al. A homozygous deletion in GRID2 causes a human phenotype with cerebellar ataxia and atrophy. J Child Neurol 2013; 28: 926–32.

Van Bon BWM, Balciuniene J, Fruhman G, Nagamani SCS, Broome DL, Cameron E, et al. The phenotype of recurrent 10q22q23 deletions and duplications. Eur J Hum Genet 2011; 19: 400–8.

Van Schil K, Meire F, Karlstetter M, Bauwens M, Verdin H, Coppieters F, Scheiffert E, Van Nechel C, Langmann T, Deconinck N, De Baere E. Early-onset autosomal recessive cerebellar ataxia associated with retinal dystrophy: new human hotfoot phenotype caused by homozygous GRID2 deletion. Genet Med. 2015 Apr;17(4):291–9.

Wilkerson JR, Albanesi JP, Huber KM. Roles for Arc in metabotropic glutamate receptor-dependent LTD and synapse elimination: Implications in health and disease. Semin Cell Dev Biol 2018; 77: 51–62.

Wu G, Robertson DH, Brooks CL 3rd, Vieth M. Detailed analysis of grid-based molecular docking: A case study of CDOCKER-A CHARMm-based MD docking algorithm. J Comput Chem. 2003 Oct;24(13):1549–62. doi: 10.1002/jcc.10306. PMID: 12925999.

Xing L, Larsen RS, Bjorklund GR, Li X, Wu Y, Philpot BD, Snider WD, Newbern JM. Layer specific and general requirements for ERK/MAPK signaling in the developing neocortex. Elife. 2016 Feb 5;5:e11123.

Yadav R, Gupta SC, Hillman BG, Bhatt JM, Stairs DJ, Dravid SM. Deletion of glutamate delta-1 receptor in mouse leads to aberrant emotional and social behaviors. PloS One 2012; 7: e32969.

Yadav R, Hillman BG, Gupta SC, Suryavanshi P, Bhatt JM, Pavuluri R, et al. Deletion of glutamate delta-1 receptor in mouse leads to enhanced working memory and deficit in fear conditioning. PloS One 2013; 8: e60785.

Yap CC, Murate M, Kishigami S, Muto Y, Kishida H, Hashikawa T, Yano R. Adaptor protein complex-4 (AP-4) is expressed in the central nervous system neurons and interacts with glutamate receptor delta2. Mol Cell Neurosci. 2003 Oct;24(2):283–95.

Yuzaki M, Aricescu AR. A GluD Coming-Of-Age Story. [Review]. Trends Neurosci 2017; 40: 138–50.

Zagrebelsky M, Holz A, Dechant G, Barde YA, Bonhoeffer T, Korte M. The p75 neurotrophin receptor negatively modulates dendrite complexity and spine density in hippocampal neurons. J Neurosci. 2005 Oct 26;25(43):9989–99.

Zhang T, Hou L, Chen DT, McMahon FJ, Wang JC, Rice JP. Exome sequencing of a large family identifies potential candidate genes contributing risk to bipolar disorder. Gene 2018; 645: 119–23.

